# Protocol for the PEACH in Asia Study: A Prospective Multinational Multicenter Observational Study on the Epidemiology of Severe Critical Events in Pediatric Anesthesia in Asia

**DOI:** 10.1101/2022.11.13.22282262

**Authors:** Soichiro Obara, Choon Looi Bong, Norifumi Kuratani, Zehra Serpil Ustalar Ozgen, Mahin Seyedhejazi, Shemila Abbasi, Ekta Rai, Elsa Varghese, Evangeline K Villa, Teresita A Batanes, Andi Ade W Ramlan, Ina Ismiarti Shariffuddin, Rufinah Teo, Patcharee Sriswasdi, Pheakdey Nhoung, Vivian Yuen, Hyo-Jin Byon, Josephine S K Tan, Asian Society of Paediatric Anaesthesiologists (ASPA) research special interest group

**Author notes:** Corresponding author: Soichiro Obara, Teikyo University Graduate School of Public Health, Tokyo, Japan, Asian Society of Paediatric Anaesthesiologists Research Special Interest Group E-mail-1, E-mail-2.

## Abstract

**Background:** Despite significant advancements in pediatric anesthesia training and guidelines, the incidence of severe critical events in children undergoing anesthesia in Asia remains under-researched. This knowledge gap is particularly pressing given the rare but life-threatening nature of such complications. While studies from high-income countries report a decline in pediatric anesthesia-related mortality, similar data from developing regions, including Asia, are sparse and suggest higher risks.

**Objective:** The Peri-anesthetic Morbidity in Children in Asia (PEACH in Asia) study aims to provide a comprehensive assessment of the incidence and risk factors associated with severe critical events in pediatric anesthesia across Asia. This multinational, multicenter, prospective observational study seeks to enhance understanding of pediatric anesthesia-related risks and variability in practices within the region.

**Methods:** The study will enroll children aged birth to 15 years undergoing elective or urgent diagnostic or surgical procedures requiring sedation or general anesthesia, with or without regional analgesia. Data collection will focus on severe critical events occurring during and up to 60 minutes post-anesthesia, including laryngospasm, bronchospasm, pulmonary aspiration, drug errors, anaphylaxis, cardiovascular instability, neurological damage, cardiac arrest, and post-anesthetic stridor. The study will also capture patient demographics, medical history, and procedural details to identify potential risk factors.

**Results:** The pilot phase of the PEACH in Asia study, conducted from May to June 2023, included 330 patients from ten institutions across nine countries. Preliminary findings revealed a severe critical event incidence rate of 12.4% [95% CI: 9.2-16.4]. Based on these results, the main study plans to recruit approximately 10,958 children across 15 to 30 Asian countries to achieve robust statistical power and refine risk factor identification.

**Conclusion:** The PEACH in Asia study represents a critical step towards understanding and mitigating the risks associated with pediatric anesthesia in Asia. By providing data-driven insights into the incidence of severe critical events and regional variations in anesthesia practices, this study aims to inform and improve pediatric anesthesia protocols across the continent.

## 1. General Information

### 1.1 Steering Committee

**Asian Society of Pediatric Anesthesiologists Research Interest Group**

**Soichiro Obara** (Teikyo University Graduate School of Public Health, Tokyo, Japan & Tokyo Metropolitan Ohtsuka Hospital) (principal investigator, Tokyo, Japan)

**Choon Looi Bong** (KK Women’s and Children’s Hospital, Singapore, Sinagpore) (co-principal investigator)

**Norifumi Kuratani** (Saitama Children’s Medical Center, Saitama, Japan) (co-principal investigator)

**Zehra Serpil Ustalar Ozgan** (Acibadem Altunizade Hospital, Acibadem Mehmet Ali Aydinlar University, Istanbul, Turkey)

**Mahin Seyedhejazi** (Tabriz University of Medical Sciences, Tabriz, Iran)

**Shemila Abbasi** (Aga Khan University, Karachi, Pakistan)

**Elsa Varghese** (Kasturba Medical College & Hospital, Manipal University, Manipal, India) **Evangeline K Villa** (Philippine General Hospital, University of the Philippines College of Medicine, Manila, Philippines)

**Teresita A Batanes** (UERM Memorial Medical Center, Quezon City, Metro Manila, Philippines) **Andi Ade W Ramlan** (Dr. Cipto Mangunkusumo Hospital, Universitas Indonesia, Jakarta, Indonesia)

**Ina Ismiarti Shariffuddin** (University of Malaya, Kuala Lumpur, Malaysia)

**Rufinah Teo** (Universiti Kebangsaan Malaysia, Bangi, Malaysia)

**Patcharee Sriswasdi** (Siriraj Hospital, Mahidol University, Bangkok, Thailand) **Pheakdey Nhoung** (National Pediatric Hospital, Phnom Penh, Cambodia) **Vivian Yuen** (Hong Kong Children’s Hospital, Hong Kong, China)

**Hyo-Jin Byon** (Yonsei University College of Medicine, Seoul, Republic of Korea)

**Josephine S K Tan** (KK Women’s and Children’s Hospital, Singapore, Singapore)

#### Sponsorship

The PEACH in Asia study is sponsored by the Asian Society of Paediatric Anaesthesiologists (ASPA) research special interest group. The aim of the ASPA research special interest group is to provide an infrastructure for clinical research in the fields of pediatric anesthesia mainly by international collaborative studies.

The Steering Committee of this study, PEACH in Asia study, can be contacted via: **E-mail-1: E-mail-2, Webpage: (under construction as of August 16th, 2024)**

#### Endorsement

The PEACH in Asia study has been endorsed by the following societies/organizations: - **Asian Society of Pediatric Anesthesiologists (ASPA)**

### 1.2 Summary of the PEACH in Asia study

Despite advancements in pediatric anesthesia training programs and the development of various guidelines for pediatric anesthesia services, the incidence of severe critical events in children remains poorly understood in Asia. Given that major life-threatening complications associated with general or regional anesthesia are rare, there is a pressing need for a large-scale, multinational, multicenter study to provide accurate statistical estimates and identify risk factors for such severe critical events. To address this gap, the international prospective observational multicenter study, **<u>Peri-anesthetic Morbidity in Children in Asia (PEACH in</u> <u>Asia)</u>**, has been designed. This study aims to determine the incidence and potential risk factors for severe critical events in children undergoing anesthesia across Asia. The findings from this research will serve as a valuable guide for developing and implementing data-driven pediatric surgical and anesthesia protocols throughout the region.

## 2. Introduction and Background Information

### 2.1 Summary of Findings from Clinical Studies Related to the PEACH in Asia Study

Numerous pediatric studies conducted in high-income countries have demonstrated a decline in pediatric anesthesia-related mortality to approximately 1-5 per 10,000 anesthetic procedures (0.01-0.05%), despite variations in study methodologies [1–7]. In contrast, data from developing countries are more limited, yet available evidence suggests that pediatric anesthesia-related mortality in these regions may be two to three times higher than in developed countries [6,7]. The incidence of serious adverse events related to pediatric anesthesia in developed countries remains around 2-8% [6–10]. The recent Anesthesia Practice in Children Observational Trial (APRICOT) highlighted considerable variability in anesthesia practices across 33 European countries, revealing a high overall incidence of 5.2% for perioperative severe critical events [11]. Similarly, data from Asian countries show comparable rates of anesthesia-related adverse events: 4.6% among infants in Thailand [12], 3.3% across all pediatric patients in Singapore [13], and 8.9% among patients aged 15 years or younger in India [14].

While the rates of pediatric anesthesia-related cardiac arrests and serious adverse events differ by region, certain high-risk factors are consistently identified across both developed and developing countries. These factors include: being younger than 1 year of age, having an American Society of Anesthesiologists (ASA) Physical Status Classification of III-V, and undergoing emergency surgery [6,7]. Notably, up to 75% of anesthetic-related cardiac arrests and critical incidents are deemed preventable [1–3, 6, 7]. Increasing awareness and understanding of common perioperative pediatric life support scenarios is essential for improving safety and quality in this field [15]. In the UK and North America, simulation-based training courses such as Managing Emergencies in Pediatric Anesthesia (MEPA) have been introduced to enhance practitioners’ readiness for anesthetic emergencies in children [16]. However, the efficacy of such simulation training in improving outcomes remains an area of ongoing investigation [16–19].

In response to these challenges, the Asian Society of Pediatric Anesthesiologists (ASPA) initiated Pediatric Perioperative Life Support (PPLS) courses in 2014, which have been conducted by experienced pediatric anesthesiologists from various Asian countries. Since then, ASPA has provided instructors and training materials, organized workshops, and developed programs in countries including the Maldives, Malaysia, Mauritius, Cambodia, Lao PDR, India, Philippines, Sri Lanka, China, Thailand, Turkey, Indonesia, and Nepal, in chronological order. Detailed reports of each course are published, regularly updated, and accessible on the ASPA official website [20].

As with many international scientific meetings, conferences, and educational courses, nearly all ASPA Pediatric Perioperative Life Support (PPLS) courses have been suspended due to the ongoing COVID-19 pandemic. Consequently, the pandemic has necessitated a shift in educational approaches, similarly impacting anesthesiology training [21–23]. For instance, in July 2020, the ASPA initiated a series of online educational lectures delivered by members of the ASPA Educational Committee. This initiative has continued on a monthly basis since July 2021, with financial support from the World Federation of Societies of Anesthesiologists (WFSA) [24].

Despite the advancement of structured programs for pediatric anesthesia training, there remains a paucity of data on whether ASPA’s PPLS courses have effectively reduced morbidity and mortality in pediatric perioperative care across various regions in Asia. As demonstrated by other studies evaluating the impact of educational courses on anesthesia practices [25,26], it is crucial for ASPA to assess the practical outcomes of our PPLS courses beyond their theoretical contributions to patient safety. Metrics such as 24-hour perioperative and anesthesia-related morbidity and mortality rates are vital indicators of access to and safety within pediatric surgical care [6,7,27]. Consequently, as a continuation of international collaborative research, ASPA plans to undertake a joint clinical research project aimed at evaluating pediatric perioperative morbidity and mortality rates across Asian countries and regions. This study will employ inclusive definitions and straightforward data collection methods. Following the model of the European prospective multicenter observational study [11], we propose a prospective observational multicenter cohort study designed to investigate the incidence of serious adverse events among neonates and children undergoing anesthesia, while also elucidating regional differences in pediatric anesthesia practices within Asia.

The study will encompass all children from birth to 15 years of age scheduled for elective or urgent diagnostic or surgical procedures under sedation or general anesthesia, with or without regional analgesia. This cohort represents the denominator for calculating the incidence of severe critical events, which is the primary objective of the study.

Anesthesiologists will document occurrences of selected severe critical events during and up to 60 minutes after anesthesia or sedation, provided these events necessitate immediate intervention. These severe critical incidents include: laryngospasm, bronchospasm, pulmonary aspiration, drug error, anaphylaxis, cardiovascular instability, neurological damage, peri-anesthetic cardiac arrest, and post-anesthetic stridor. Additionally, pertinent aspects of the child’s medical and family history will be recorded.

The PEACH in Asia pilot study was conducted from May 1 to June 30, 2023, at ten institutions across nine countries or regions in Asia, namely Türkiye, Pakistan, India, Indonesia, Singapore, Malaysia, the Philippines, Hong Kong, and Japan (with two institutions in Japan). The participating institutions were selected based on their affiliation with members of the ASPA research special interest group. The pilot study successfully enrolled 330 patients across nine institutions in these nine countries or regions. However, one institution in Japan withdrew from the study due to the substantial burden associated with data collection and entry into the electronic case report form (e-CRF), compounded by the research coordinator’s clinical workload during the three-day recruitment period.

Among the 330 enrolled patients, 36 children (10.9%) experienced severe critical events, with a total of 41 severe critical events occurring during or immediately following anesthesia or sedation. This corresponds to an incidence rate of 12.4% [95% CI: 9.2-16.4]. Based on this observed incidence rate, the estimated sample size for the main study was calculated to be 10,958, using a standard binomial assumption to achieve a 95% confidence level. Consequently, we plan to recruit at least 10,958 children over a two-week period, including weekends and after-hours, across 15 to 30 Asian countries or regions represented at the Asian Society of Pediatric Anesthesiologists Council or located within geographical Asia. The recruitment period will be determined by each site and is scheduled to commence in August, 2024. The final inclusion date will be set by the Study Steering Committee, depending on the recruitment rate.

Participating institutions will be provided with data acquisition sheets for anonymous and standardized recording of patient parameters, which will be used to complete the electronic case report form (e-CRF).

Data analysis will employ descriptive statistics to summarize patient demographics and the incidence of severe critical events. Additionally, exploratory statistical analyses, such as hierarchical logistic regression or relative risk models, will be conducted to identify independent risk factors for severe critical events, with results reported as adjusted odds ratios or relative risks.

### 2.2 Compliance with the Study Protocol, Good Clinical Practice Standards, and Applicable Regulatory Requirements

This study will be conducted in strict adherence to the approved protocol, which has been reviewed and sanctioned by the Institutional Review Board (IRB). It will also conform to Good Clinical Practice (GCP) standards throughout its execution. Any substantial amendments to the protocol will not be implemented without prior review and approval from the IRB. However, given the observational nature of this study, such amendments are considered highly unlikely.

In addition, the study will comply with the specific regulatory requirements of each participating country. These regulations will be carefully followed to ensure that the study is conducted ethically and in accordance with local legal and professional standards.

## 3. Trial Objective and Purpose

**The objectives of the PEACH in Asia study are as follows:**

1. **To determine the incidence of severe critical events in children undergoing anesthesia across Asia.**
2. **To characterize the variability in pediatric anesthesia practices throughout the region.**
3. **To investigate the potential impact of this variability on the occurrence of severe critical events, including laryngospasm, bronchospasm, pulmonary aspiration, drug error, anaphylaxis, cardiovascular instability, neurological damage, peri-anesthetic cardiac arrest, and post-anesthetic stridor.**

In contrast to Europe and North America, there is a notable lack of data concerning morbidity and mortality associated with pediatric peri-anesthetic care in Asia. This study represents the first comprehensive effort to examine the incidence of severe critical events in children undergoing anesthesia across diverse Asian settings. The insights gained from this research will serve as crucial indicators of access to and safety within pediatric surgical care throughout Asia.

## 4. Study Design

### 4.1 Type/ Design of Study

▪ Study Type: Observational
▪ Observational Model: Cohort
▪ Time Perspective: Prospective
▪ Official Title: PEACH in Asia: PEri-Anesthetic Morbidity in CHildren in Asia – A Prospective Multinational Multicenter Observational Study to Investigate the Epidemiology of Severe Critical Events in Pediatric Anesthesia in Asia
▪ Planned Study Start Date: August 31, 2024
▪ Projected Primary Completion Date: March 31, 2026
▪ Projected Study Completion Date: September 30, 2026

The PEACH in Asia study is a prospective, descriptive observational study that collects objective data as part of routine anesthetic care. As an observational trial, it does not involve any interventions; thus, it is neither double-blind nor placebo-controlled. The study focuses on standardized data acquisition without introducing specific procedural changes.

### 4.2 Outcome Measures

#### 4.2.1 Primary Endpoint

**Incidence of Severe Critical Events [Time Frame: Children will be monitored throughout their anesthesia procedure and up to 60 minutes post-anesthesia or sedation]**

The primary endpoint is the incidence of severe critical events, defined as any incident occurring during or up to 60 minutes after anesthesia or sedation (including in the Post-Anesthesia Care Unit [PACU]) that requires immediate intervention and may lead to major disability and/or death. The specific severe critical events include:

1. **Laryngospasm**
2. **Bronchospasm**
3. **Pulmonary aspiration**
4. **Drug error**
5. **Anaphylaxis**
6. **Cardiovascular instability**
7. **Neurological damage**
8. **Peri-anesthetic cardiac arrest**
9. **Post-anesthetic stridor**

#### 4.2.2 Secondary Endpoint

**Risk Factors for Severe Critical Events [Time Frame: Children will be monitored throughout their anesthesia procedure and up to 60 minutes post-anesthesia or sedation]**

The secondary endpoint involves identifying risk factors associated with the occurrence of severe critical events. This will be achieved by collecting comprehensive data on:

1. Social and demographic information of the patients
2. Family and child’s medical history
3. Presence of co-morbidities
4. Details of the anesthesia procedure
5. Whether the case was elective or emergency
6. Level of experience of the anesthesiologist
7. Postoperative prescriptions

### 4.3 Data Collection

The following data will be collected for each patient:

#### 4.3.1 Demographic Data

- Age
- History of prematurity
- Gender
- Weight
- Ethnicity
- ASA Physical Status Classification

#### 4.3.2 Medical History

- Presence of current upper respiratory infection (within the past 2 weeks)
- History of wheezing, chronic dry cough at night, or eczema within the last 12 months
- Known allergies and atopy
- Fever exceeding 38.5°C in the last 24 hours
- Family history of smoking
- Medication and/or Natural or Homeopathic Products

#### 4.3.3 Parameters Related to the Anesthesia Team and the Procedure

- Details of the pre-anesthetic consultation
- Information about the anesthesia team
- Indication for the procedure (surgical or non-surgical)
- Timing of the procedure (elective or emergency; during operating room hours or outside regular hours)
- Ambulatory or inpatient status **Definition of Emergency Procedure:** A non-elective procedure performed when the patient’s life or well-being is in direct jeopardy. Conversely, a procedure scheduled in advance without medical emergency or urgency is considered elective.

#### 4.3.4 Type of Premedication and Route of Administration

- Details of premedication used and its route of administration
- Presence of parents at induction

#### 4.3.5 Anesthesia Management

- Type of monitoring employed
- Variables related to induction (type, sequence, and drugs used)
- Maintenance phase (drugs administered, use of nitrous oxide)
- Any additional regional analgesia (technique, test dose, drugs used)
- Airway management (type used, difficulties encountered, use of cuffed tubes, monitoring and technique for removal on awakening)
- Type of ventilation used
- Fluid management (maintenance, compensation for fluid losses, blood transfusion, coagulation factors)
- Duration of the procedure (defined as wheel-in/wheel-out, indicating when the patient enters or leaves the operating room, or when the anesthesiologist begins care if the procedure is performed outside the operating room)

#### 4.3.6 Recording of Severe Critical Events Occurring During Anesthesia and Up to 60 Minutes Afterwards Defined as It Follows

The following definitions, mostly based on validated or published studies in the field of pediatric anesthesia will be used in the PEACH in Asia study:

1. Laryngospasm: is defined either as complete airway obstruction associated with rigidity of the abdominal and chest walls and leading to unsuccessful child’s ventilation, or glottic closure associated with chest movement but silent unsuccessful child’s respiratory efforts and assisted ventilation, unrelieved in both situations with simple jaw thrust and CPAP maneuvers and requiring the administration of medication (propofol, suxamethonium etc.) and/or tracheal intubation.
2. Bronchospasm: is defined as an increased respiratory effort, especially during expiration, and wheeze on auscultation. If the patient is ventilated, bronchospasm may also be considered if a significant increase in peak inspiratory pressure (under volume controlled ventilation) or significant decrease in tidal volume (under pressure controlled ventilation) are observed. In all cases, any episode of airway constriction requiring the administration of a bronchodilator will be recorded.
3. Pulmonary aspiration: is defined as the presence of any non-respiratory secretions (bilious or particulate) in the airway as evidenced by laryngoscopy, suctioning, or bronchoscopy. In a situation where there was suspicion of pulmonary aspiration but no positive aspiration of non-respiratory secretions, new clinical and/or chest X-ray signs consistent with aspiration are accepted as evidence for it (e.g., new wheeze or crackles in the chest after regurgitation or vomiting incident).
4. Drug error: is defined as the administration of a wrong drug, or a wrong dose given by any route, or a wrong site of administration, that has led to either respiratory/cardiac/neurological consequence or to an unplanned admission to the intensive care unit or prolonged hospitalization.
5. Anaphylaxis: is defined by the occurrence of any suspected IgE or non-IgE mediated severe allergic reaction leading to cardiovascular instability and/or severe bronchospasm and requiring immediate resuscitation (fluid resuscitation and epinephrine).
6. Cardiovascular instability: is defined by the occurrence of either one of the following:

a) cardiac arrhythmia defined as ECG evidence of cardiac rhythm disturbance considered by clinical staff to be severe enough to require treatment (e.g. anti- arrhythmic agents, vasoactive agents, intravenous fluid, etc.). This includes arrhythmias occurring following regional analgesia and requiring intervention. For example: bradycardia requiring atropine, supraventricular tachycardia, atrial or ventricular tachyarrhythmia, torsade de Pointe, etc.
b) hypotension defined as a drop in blood pressure requiring intervention by the anesthesiologist (fluid resuscitation and/or the administration of vasoactive drugs)
c) bleeding resulting in hypotension and necessitating unanticipated and unpredicted blood transfusion
d) cardiovascular instability despite anticipated bleeding and transfusion (e.g.: liver transplant, scoliosis…)
7. Neurological damage: is defined in case of regional anesthesia by the occurrence of nerve injury or spinal cord insult or seizure requiring resuscitation. In case of general anesthesia, any episode of seizure, pressures sore, episodes of loss of vision or new onset of central neurological impairment. This includes peripheral nerve injury following positioning (ulnar nerve, external popliteal nerve) or puncture (median or ulnar nerve).
8. Peri-operative cardiac arrest: is defined as cessation of circulation (e.g. pulseless electric activity, asystole, ventricular fibrillation/tachycardia) requiring open or closed chest compressions, or resulting in death, while the patient is in the care of the anesthetic team. Three advocators, among the local anesthetic team, will determine the anesthetic responsibility for cardiac arrest.
9. Post-operative Stridor: is defined as a severe inspiratory flow limitation with sternal retraction, intrathoracic pressure swing, and potentially cyanosis occurring in the PACU and necessitating the administration of oxygen, intravenous steroids and/or epinephrine (nebulization) or tracheal intubation.

Postoperative Data:

- Information on the patient’s transfer and the health providers in charge will be recorded.
- In addition to the severe critical events previously listed, it will be noted whether the child received supplemental oxygen.

At the conclusion of the study period, each participating institution will submit an "End of Study Reporting Form," which will include:

- The total number of surgical procedures performed under anesthetic care during the study period
- The total number of patients who were screened but excluded from the study

Additionally, each institution will provide a Screening Failure Tracking Form at the end of the study period. This form will facilitate the analysis of reasons for exclusion from the study, such as:

- Refusal to sign informed consent (if applicable)
- Changes to the operation list
- Last-minute cancellations
- Other relevant reasons

### 4.4 Description of Measures Taken to Minimize/Avoid Bias

The PEACH in Asia study is inherently descriptive and relies solely on objective data collected as part of routine anesthetic care. To mitigate potential biases, the following measures have been implemented:

#### 4.4.1 Inclusivity and Non-Interference

All patients under 16 years of age will be included in each participating instituion. Participation in the study will not alter the clinical care provided to the patients, as the study is observational in nature.

#### 4.4.2 Data Anonymization and Secure Collection

- All data will be anonymized and collected via a secure website. Given the observational nature of the study, it is anticipated that local Institutional Review Boards (IRBs) may waive the requirement for written parental consent, as detailed in the ethics section.

#### 4.4.3 Coordination and Oversight

To ensure the adequacy and accuracy of data collection, national and local coordinators will be appointed with specific responsibilities:

**National Coordinators:** Appointed by the Steering Committee, national coordinators will:

- Identify and liaise with local coordinators at participating institutions.
- Assist with the translation of study documentation as needed.
- Ensure that necessary country or regional regulatory approvals are secured prior to the study’s start.
- Maintain effective communication with participating sites within their nation. **Local Coordinators:** Local coordinators at individual institutions will:
- Provide leadership and oversight for the study within their institution.
- Ensure that all relevant regulatory approvals are obtained for their institution.
- Facilitate adequate training for all relevant staff prior to the commencement of data collection.
- Supervise daily data collection and provide assistance with any issues that arise.
- Guarantee the integrity and quality of the data collected.
- Maintain communication with the national coordinator. These measures are designed to uphold the integrity of the study, ensure accurate data collection, and minimize any potential biases.

### 4.5 Expected Duration of Subject Participation and Description of Study Periods

#### 4.5.1 During Anesthesia and Immediate Post-Anesthesia Monitoring

- Patients will be observed throughout the duration of their anesthesia management and up to 60 minutes immediately following the procedure. This phase includes monitoring for any severe critical events that occur during or shortly after anesthesia.

#### 4.5.2 Data Collection Timeline

- Initial Data Acquisition: Data will be collected on the day of the anesthesia management, covering both the anesthesia procedure and immediate post-anesthesia period.

### 4.6 Stopping Rules or Discontinuation Criteria

For the PEACH in Asia study, the following criteria apply to the discontinuation of individual subjects, parts of the study, or the entire study:

#### 4.6.1 Withdrawal of Consent

Subjects may withdraw from the study at any time. Upon withdrawal, data collected up to the point of withdrawal will be retained for analysis unless otherwise specified by the subject.

#### 4.6.2 Discontinuation Criteria for Individual Subjects

There are no specific discontinuation criteria for individual subjects beyond voluntary withdrawal of consent. If a subject experiences a severe adverse event, their participation will continue as required to collect pertinent data, but their involvement in further study activities will be at their discretion.

#### 4.6.3 Discontinuation Criteria for Parts of the Study

No specific discontinuation criteria are defined for parts of the study. However, any issues impacting the integrity of data collection or the safety of participants will be addressed promptly. If significant issues arise, relevant adjustments or modifications will be made as necessary.

#### 4.6.4 Discontinuation Criteria for the Entire Study

There are no predefined criteria for discontinuing the entire study. The study will continue unless terminated due to unforeseen circumstances that jeopardize participant safety, compliance with regulatory requirements, or if the study objectives are deemed unattainable. Any decision to halt the study will be made by the Steering Committee in conjunction with appropriate regulatory bodies and will be communicated to all participating institutions.

This framework ensures that participant safety and data integrity are maintained throughout the study, while providing flexibility to address any issues that may arise.

### 4.7 Pilot Testing

#### 4.7.1 Purpose

A pilot study will be conducted prior to the full-scale implementation of the main trial. The purpose of this pilot study is to assess and refine all study procedures on a smaller scale. This includes evaluating the selection and enrollment of eligible participants, recording the required data, implementing supervision systems, ensuring quality control, and processing data. The pilot study aims to identify and address any issues in these processes before the main trial commences.

#### 4.7.2 Design of the Pilot Study

The pilot study will closely mirror the design and procedures of the main trial, ensuring consistency and relevance. The selected population for the pilot study will be representative of the trial population but will not overlap with participants in the main trial. This approach avoids the need to revisit the same individuals for data collection in the main trial.

Key aspects of the pilot study will include:

- **Testing Procedures:** Every aspect of the main trial’s processes will be tested, including participant selection, data collection methods, and data management systems.
- **Data Handling:** Pilot data will be entered into the data management system, cleaned, and analyzed. This step is crucial for verifying the functionality and effectiveness of data processing and quality control systems.

Results from the pilot study may be published as “Feasibility and Pilot Study of the PEACH in Asia Study.”

## 5 Selection and Withdrawal of Subjects

### 5.1 Subject Inclusion/ Exclusion Criteria

#### 5.1.1 Eligibility Criteria

- Ages Eligible for Study: Up to 15 Years (Child)
- Sexes Eligible for Study: All
- Accepts Healthy Volunteers: No
- Sampling Method: Non-Probability Sample

#### 5.1.2 Study Population

The study will include all children aged less than 16 years who are scheduled for an elective, urgent, or emergency diagnostic or surgical procedure under sedation or general anesthesia, with or without regional analgesia.

**Inclusion Criteria:**

✓ Age: from birth to 15 years included
✓ All children admitted for an inpatient or outpatient procedure under general anesthesia with or without regional analgesia or under regional anesthesia alone. This includes all kind of surgeries and procedures requiring anesthesia and analgesia to be performed such as central venous access, burn care, cast, etc.
✓ Children admitted for a diagnostic procedure under sedation (performed by an anesthesiologist) or general anesthesia (such as endoscopy, radiology (CT-scan, MRI), cardiac catheterization and electrophysiology, PET-scan, radiotherapy, lumbar and bone marrow puncture, biopsies), diagnostic procedure under general anesthesia (such as endoscopy, radiology)
✓ Children admitted for urgent or emergency procedure performed in- or out-of-hours

**Exclusion Criteria:**

✗ Children admitted directly from the intensive care units to the operating rooms

✗ Anesthesia procedures in the neonatal or pediatric intensive care units

✗ Age: All children aged ≥ 16 years

### 5.2 Subject Withdrawal Criteria

Apart from withdrawal of consent, there are no specific criteria for withdrawing subjects from the study.

Withdrawal of Consent:

- If a subject withdraws consent, all data collection for that individual will cease immediately. The Case Report Form (CRF) and the electronic Case Report Form (e-CRF) for the withdrawn subject will be left incomplete.
- Once consent is withdrawn, no further follow-up will be conducted for the subject.
- Subjects who withdraw consent will not be replaced, and their data will not be included in the final analysis.

## 6 Statistics

### 6.1 Sample Size Calculation for the Main Study

The following formula was used to estimate the required sample size (***n***) for the main study:

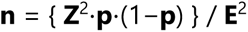

where:

- **Z** is the Z-value corresponding to the desired confidence level (1.96 for 95% confidence, 2.576 for 99% confidence).
- **p** is the anticipated incidence rate, which is derived from the SCE rate of the pilot study.
- **E** is the allowable margin of error, which can be calculated as the half of the confidence interval (CI) width when the CI width is known.

Using the APRICOT study’s reported SCE rate of 5.2% [95% CI: 5.0, 5.5] [6], the CI width was approximately 10% (±5%) of the incidence rate. Thus, **E** is set to 5% of the SCE rate, derived from the pilot study results.

The PEACH in Asia pilot study was conducted from May 1 to June 30, 2023, at ten institutions across nine countries or regions in Asia, namely Türkiye, Pakistan, India, Indonesia, Singapore, Malaysia, the Philippines, Hong Kong, and Japan (with two institutions in Japan). The participating institutions were selected based on their affiliation with members of the ASPA research special interest group. The pilot study successfully enrolled 330 patients across nine institutions in these nine countries or regions. However, one institution in Japan withdrew from the study due to the substantial burden associated with data collection and entry into the electronic case report form (e-CRF), compounded by the research coordinator’s clinical workload during the three-day recruitment period.

Among the 330 enrolled patients, 36 children (10.9%) experienced severe critical events, with a total of 41 severe critical events occurring during or immediately following anesthesia or sedation. This corresponds to an incidence rate of 12.4% [95% CI: 9.2-16.4]. Based on this observed incidence rate, the estimated sample size for the main study was calculated to be 10,958, using a standard binomial assumption to achieve a 95% confidence level.

### 6.2 Participating Institutions

The PEACH in Asia study aims to engage a broad range of participating institutions across Asian countries and regions represented by the Asian Society of Pediatric Anesthesiologists (ASPA) council, as well as other areas within Asia. Recruitment will occur over a two-week period, which will include weekends and after-hours, and each participating site will select its specific recruitment window within 2024-2025.

#### Eligibility for Participation

- **Institution Type:** Both private and public institutions, whether academic, regional, or referral institutions, are eligible to participate.
- **Pre-Study Requirements:** Prior to enrolling the first patient, each participating institution must complete a pre-study questionnaire. This questionnaire will capture information about the type and size of the hospital and relevant parameters concerning local standards of care.

#### Coordination

- **Local Coordinators:** Each participating institution will designate a local coordinator responsible for managing the study within their institution.
- **National Coordinators:** A national coordinator will oversee the recruitment process within each country. They will provide support to participating institutions, address any questions, and monitor recruitment progress. For detailed information on the responsibilities of national and local coordinators, please refer to the earlier section on coordination. All institutions are encouraged to participate in this collaborative project to contribute to the study’s objectives and enhance the understanding of pediatric anesthesia practices across Asia.

### 6.3 Analytical Method

The PEACH in Asia study is a descriptive observational study, and the analysis will primarily involve descriptive statistical methods. The main outcome measure, the incidence of severe critical events, will be reported along with its 95% confidence interval.

#### Data Collection and Analysis

- **Data Source:** All data collected during the study will be part of routine clinical care. There will be no alteration to standard clinical practices.
- **Categorical Variables:** These will be reported as proportions. Comparisons between categorical variables will be conducted using chi-square tests or Fisher’s exact tests, as appropriate.
- **Continuous Variables:** These will be described using means and standard deviations if normally distributed. If not normally distributed, data will be presented as medians and interquartile ranges. Comparisons of continuous variables will be performed using one-way ANOVA or Mann-Whitney U tests, depending on the distribution of the data.
- **Univariate and Multivariate Analysis:** Univariate analysis will initially assess factors associated with the study endpoints. Multivariate analysis will employ hierarchical logistic regression or relative risk models to identify independent risk factors. Variables included in the multivariate models will be selected based on p-values from univariate tests (p<0.05) and clinical relevance.
- **Multicollinearity:** Multicollinearity will be assessed and addressed. A stepwise elimination process will be used to refine the model, considering medically plausible interactions between variables. Results will be reported as adjusted odds ratios or relative risks with 95% confidence intervals.
- **Final Analysis:** A comprehensive final analysis will be conducted at the conclusion of the study. Given the descriptive nature of the trial and the absence of interventions altering routine care, no interim analyses are planned. **Procedures for Accounting for Missing, Unused, and Spurious Data**
- **Handling of Missing Data:** All patients enrolled in the study will be included in the data analysis, irrespective of the completeness of their electronic Case Report Form (e-CRF) data.
- **Exclusion of Data:** Unused and spurious data will be identified and excluded from the analysis upon detection. This will ensure that only valid and reliable data contribute to the study’s findings. **Selection of Subjects for Analysis** All subjects who are included in the study will be part of the data analyses. This includes any patient who has been enrolled, regardless of the completeness or quality of the data collected.

## 7 Quality Assurance and Quality Control

The sponsor, ASPA research special interest group, is responsible for establishing and maintaining quality assurance and quality control systems to ensure that the PEACH in Asia study is conducted in compliance with the protocol, Good Clinical Practice (GCP) standards, and applicable regulatory requirements.

**Quality Assurance Responsibilities:**

- **Monitoring and Auditing:** ASPA research special interest group will ensure that all parties involved in the study provide direct access to trial-related sites, source data/documents, and reports for monitoring and auditing purposes. This includes access for inspections by both domestic and international regulatory authorities.
- **Data Handling:** Quality control measures will be applied throughout all stages of data handling to guarantee that the data are reliable and accurately processed.

**Agreements and Contracts:**

- **Principal Investigator Agreement:** Agreements with the principal investigator (Soichiro Obara) will be formalized in writing, either as part of the protocol or through a separate written agreement.
- **Participating Institutions:** The sponsor, ASPA research interest group, has not established specific contracts or agreements with participating institutions regarding clinical practices or activities related to the study. Given the study’s observational nature and minimal risk to patients, there is no alteration to standard clinical care. Consequently, no specific indemnity arrangements for clinical practices are required. **Indemnity and Liability:**
- **Local Indemnity:** Any issues related to the clinical care provided within participating hospitals will be covered by the local indemnity arrangements of those institutions.

## 8 Ethics Description of Ethical Considerations Relating To The Study

The PEACH in Asia study is designed as an observational study, with no interventions or modifications to the child’s routine clinical care. This approach aligns with Good Clinical Practice (GCP) standards, and therefore, the study is expected to pose minimal ethical concerns. Nonetheless, ethical approval may be required by some participating institutions, depending on local regulations.

### 8.1 Institutional Review Board (IRB) Approval

In all cases, all participating institutions must submit the study to the local Institutional Review Board (IRB) for ethical judgment and obtain document of proof that the trial has been subject to IRB review and given approval/ favorable opinion and/ or waive the consent. If informed consent is not required by the local IRB, a written waiver must be obtained from the IRB. This process should take place prior to initiation of the trial and in compliance with the applicable national regulatory requirement(s).

### 8.2 Informed Consent (only if applicable, as per local regulations)

- Elective Cases: For children undergoing elective procedures, the parents or guardians will be provided with a Patient Information Sheet during the preoperative consultation or on the ward prior to anesthesia management. Written informed consent will be obtained either during the consultation or in the holding bay or anesthetic room immediately before the procedure.
- Emergency Cases: For children undergoing emergency procedures, data will be collected during the procedure, and retrospective consent will be sought within 48 hours following admission.
- Incapacity to Consent: For parents or guardians with long-standing incapacity to consent (e.g., due to learning difficulties), a consultee (next of kin or consulting physician) should be approached to provide assent where possible. If there is insufficient time to obtain assent before the procedure, retrospective consent will be sought.

### 8.3 Documentation and Translation

- The study sponsor, ASPA research special interest group, provides templates for the Patient Information Sheet and Participant’s Informed Consent in English (only if applicable).
- Any translations or adaptations of these documents must be reviewed and approved by the Sponsor. Guidance provided by the Sponsor should be followed to ensure consistency and compliance with ethical standards.

### 8.4 Compliance and Adaptation

It is essential that all procedures related to consent and ethical considerations are conducted in accordance with applicable local regulations and the guidelines. This process is crucial for maintaining the integrity of the study and protecting the rights and welfare of participants.

## 9 Data Handling and Record Keeping

### 9.1 Data Acquisition and Anonymization

#### 9.1.1 Data Collection

- Participating institutions will receive data acquisition sheets (paper-case report forms: p- CRFs) for standardized recording of all patient parameters required for the study.
- Each local institution will be provided with a "3-digit code for the country" and a "3-digit code for the hospital" by the study sponsor.
- Each local coordinator will issue a patient identification number (PIN) in a sequential order (e.g., 001 to 999) for each participant. This will be in the format: xxx(3-digit country code) – xxx(3-digit hospital code) – xxx(3-digit individual patient number).
- A combinable anonymization correspondence table will be provided to match the unanonymized Study Subject ID with the anonymized patient identification number at each participating institution.

#### 9.1.2 Anonymization Process

- The details of the anonymization process, including parameters used and controls, must be recorded for future reference. This documentation will support review, maintenance, fine-tuning, and audits.
- The correspondence table containing patient identification numbers and anonymized IDs should be securely stored by each local coordinator in a locked location and should not be shared with national coordinators or the sponsor.

### 9.2 Data Security and Confidentiality

#### 9.2.1 Data Storage

- The data collected will be handled in an anonymous fashion. Patient names, initials, or hospital patient identification numbers will not be recorded on the p-CRF or the electronic case report form (e-CRF).
- Data will be stored securely at each local institution for the duration of the study and for a minimum of 5 years after study completion, as required by local regulations.
- Each participating institution will maintain an Investigator file, including the study protocol, IRB judgments, local investigator delegation logs, translations of informed consent forms (if applicable), and signed informed consent forms (if applicable).

#### 9.2.2 Compliance

- All personal data handling will comply with Good Clinical Practice (GCP) guidelines. The study sponsor, ASPA research interest group, retains ownership of all collected data.
- Local participating institution will ensure that data is stored and handled in a manner that maintains confidentiality and complies with applicable regulations.

### 9.3 Responsibilities and Documentation

#### 9.3.1 Local Coordinators

- Ensure that all data is anonymized appropriately and that the correspondence table is kept securely.
- Maintain all necessary documentation related to the study and ensure it is available for audits and reviews.

#### 9.3.2 Sponsor (ASPA research special interest group)

- Provide the necessary tools and guidelines for data collection and anonymization.
- Oversee the overall data management process and ensure compliance with GCP and regulatory requirements. By adhering to these procedures, the study aims to maintain the highest standards of data integrity and confidentiality, ensuring that participant information remains protected throughout the study and beyond.

## 10 Publication Policy

### 10.1 Authorship Distribution

- Authorship will be allocated based on the level of investment and contribution to the study, which includes activities such as patient recruitment, data acquisition, and analysis.
- Each participating institution that recruits a minimum of 5 patients will have the opportunity to designate one collaborator as a co-author on the publication.
- For every additional 50 patients recruited, the participating institution can designate one additional collaborator as a co-author.
- The list of collaborators will be included in the manuscript and will be traceable via PubMed.

### 10.2 Secondary Analyses and Data Use

#### 10.2.1 Proposals for Secondary Analyses

- Participating institutions can request to use their data for secondary analyses. Proposals for such analyses should be submitted to the Steering Committee, ASPA research special interest group.
- The Steering Committee will review and approve these proposals before any secondary analyses are conducted. The final papers originating from these analyses will be reviewed and approved by the Steering Committee prior to submission.

#### 10.2.2 Use of Anonymized Data

- The sponsor of the study, ASPA research special interest group, and the Asian Society of Pediatric Anesthesiologists reserve the right to use anonymized data for internal analyses and educational purposes.
- Anonymized data may be utilized to generate insights, develop educational materials, or inform future research initiatives.

### 10.3 General Guidelines

#### 10.3.1 Publication Timeline

The timeline for publication will follow the completion of data acquisition, cleaning, and analysis. The primary study findings will be published in a peer-reviewed journal.

## Supporting information

Appendix 4 (REVISED in 2024)

Appendix 6 (REVISED in 2024)

## Data Availability

The data that support the findings of this study are available from the corresponding author upon reasonable request.

## 10.3.2 Acknowledgments

In addition to co-authors, all participating institutions and their staff will be acknowledged in the publication for their contributions to the study.

## 10.3.3 Confidentiality and Data Sharing

All publications and presentations will adhere to the confidentiality agreements, ensuring that no identifiable patient information is disclosed.

This policy ensures that all participating institutions and collaborators are appropriately recognized for their contributions while maintaining the integrity and confidentiality of the study data.

## 12 Lists of Appendices

The lists of appendices:

**Appendix 1**. Patient Information Sheet in PEACH in Asia study

**Appendix 2**. Infographics of PEACH in Asia study

**Appendix 3**. Informed Consent sheet for PEACH in Asia study (required <u>only if applicable according to the local IRB</u>)

**Appendix 4**. Participating Institution Data Record Form in PEACH in Asia study

**Appendix 5**. Severe Critical Events Definitions in PEACH in Asia study

**Appendix 6**. paper Case Report Form (p-CRF) in PEACH in Asia study: REVISED on August 16^th^ 2024 according to the PEACH in Asia Pilot Study

**Appendix 7**. Correspondence Table form for PEACH in Asia study (linking Study ID and Patients’ ID at the participating institution)

**Appendix 8**. Notification of the IRB review for PEACH in Asia study, obtained in Tokyo Metropolitan Ohtsuka Hospital, Tokyo, Japan (as an international collaborative, multicenter observational study)

**Appendix 9**. Explanatory Document for Data Capturing System in the PEACH in Asia Study: UMIN-INDICE

