## Appendix 1 for "Protocol for the PEACH in Asia Study: A Prospective Multinational Multicenter Observational Study on the Epidemiology of Severe Critical Events in Pediatric Anesthesia in Asia"

### IMPORTANT PATIENT INFORMATION
### PEri-Anesthetic morbidity in CHildren in Asia (PEACH in Asia) study

**A research study is being conducted at (NAME) Hospital.**

The research is being done by (Lead Local Investigator name) from (start date) to (completion date).

**Why is this research study being done?**

To understand what complications children have during and after having an operation, procedure or examination under anesthesia or sedation.

**Why are we telling you about this study?**

All patients less than 18 years of age having an operation, procedure or examination under anesthesia or sedation in this hospital are part of the study. It is a requirement that some details pertaining to your child's clinical care are entered into a trial folder. Information from this folder will be used anonymously to see if any complications occur during or after your child's operation, procedure, or examination under anesthesia or sedation.

**Will this study affect my care while I am in hospital?**

No. Your care will not change while you are in hospital.

**Will my name or any personal details be recorded in this study?**

No. Your name and personal details will be recorded on a note for the study, but will not be recorded as part of this study. All information from the notes will be kept strictly confidential.

**Are there any risks or benefits associated with this study?**

No.

**May I withdraw from this study?**

Every patient has the right to withdraw from this study. If you would like to withdraw from the study, please contact the lead local investigator listed below.

**Who should I contact if I have any questions or concerns?**

Please contact (Name of lead local investigator) on the followings:

Telephone: ______________________________ or E-mail: ______________________________

If you have questions about your rights or welfare as a research participant, please contact the sponsor of this study, Asian Society of Paediatric Anaesthesiologists (ASPA) Research Committee through the
