## Appendix 2 for "Protocol for the PEACH in Asia Study: A Prospective Multinational Multicenter Observational Study on the Epidemiology of Severe Critical Events in Pediatric Anesthesia in Asia"

### PEri-Anesthetic morbidity in CHildren in Asia (PEACH in Asia) study

1

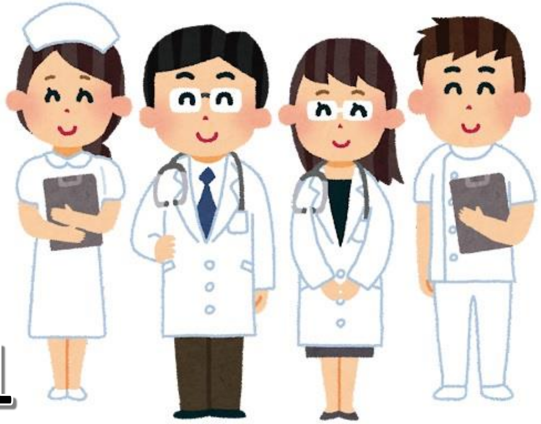

Doctors and nurses are doing a study on 'Anesthesia in Children'.

2

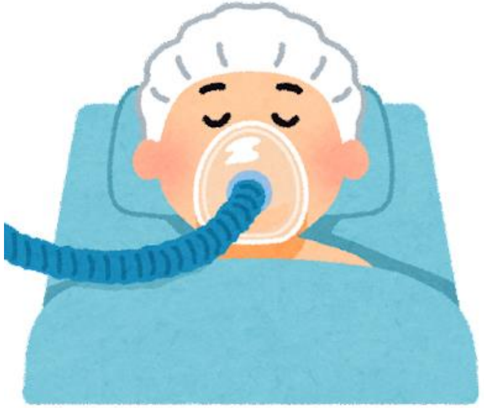

Children sometimes need anesthesia or sedation for operations, procedures or examinations.

3

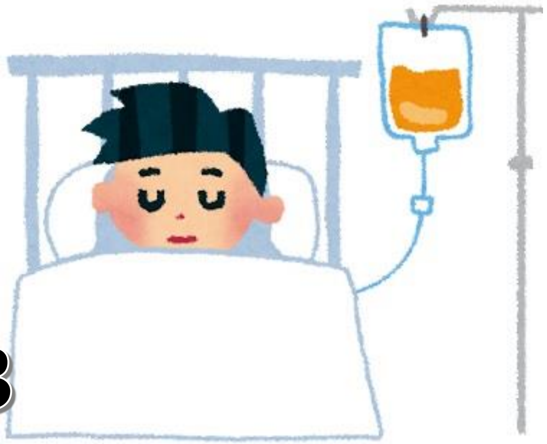

Some children have some events in hospital after anesthesia or sedation, even if not severe.

Information about your children may explain why some children have some events.

4

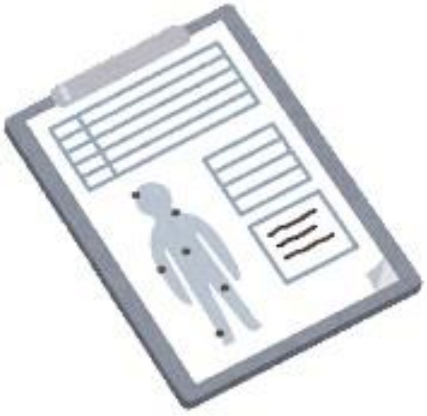

This information will be recorded from your children's hospital charts but will not include the name or address.

5

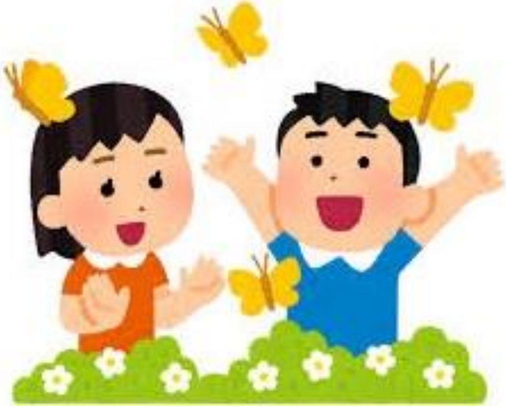

The information about your children's anesthesia or sedation may help children in the future get better.

If you want to know more about this study, you can ask the doctor in charge of this study: Dr.

___________________________

Doctors are doing a study at:
__________________ Hospital.
