## Appendix 3 for "Protocol for the PEACH in Asia Study: A Prospective Multinational Multicenter Observational Study on the Epidemiology of Severe Critical Events in Pediatric Anesthesia in Asia"

### Informed Consent Form

#### Peri-anesthetic morbidity in Children in Asia study (PEACH in Asia study)

This is an informed consent form dealing with medical research with children.

**I, __________________________, confirm that (please check box as appropriate):**

| # | Statement (please read) | Parent/ Guardian |
|---|---|---|
| 1 | I have read/ had read to me of the Important Patient Information Sheet for the above study (dated DD/MM/YYYY) and have had the opportunity to consider the information and ask questions. | ☐ |
| 2 | I understand that my child's participation in this study is voluntary and that I may withdraw them at any time without giving a reason. I understand that opting out won't affect my child's future medical care or legal rights. | ☐ |
| 3 | I give permission for researchers to look at my child's medical records to get information about their care, and to contact me as part of this research study. | ☐ |
| 4 | I give informed explicit consent to have my child's data processed as part of this research study. I am happy for information about my child related to the study being stored on password protected computer systems at your hospital and also at the Asian Society of Paediatric Anaesthesiologists (ASPA) Research Committee. This will be backed-up in a separate location to keep my child's information safe. | ☐ |
| 5 | I consent for my child to take part in this research study having been fully informed of the risks, benefits and alternatives. | ☐ |

| | |
|---|---|
| **Name of child** | **Name of Principal Investigator/ nominee taking consent** |
| **Name of Parent/ Guardian** | I, the undersigned, have taken the time to fully explain to the above parent/guardian the nature and purpose of this study in a way that they could understand. I have explained the risks involved as well as the possible benefits. I have invited them to ask questions on any aspect of the study that concerned them. |
| **Relationship of Parent / Guardian to child** | |
| **Signature (or thumb print) of Parent / Guardian** | |
| **Signature (or thumb print) of Child (if applicable)** | **Signature of researcher** |
| **Date form signed (or thumb printed) by Parent/ Guardian**<br>DD / MM / YYYY | **Date form signed by researcher**<br>DD / MM / YYYY |
