## Appendix 4 (REVISED in 2024) for "Protocol for the PEACH in Asia Study: A Prospective Multinational Multicenter Observational Study on the Epidemiology of Severe Critical Events in Pediatric Anesthesia in Asia"

### Participating Institution Data Record Form

Thank you for your valuable contribution to the PEACH in Asia study.

**Please submit your answers to the following questions using the Google Form link provided.**
https://forms.gle/sfAY5gmZ8PGQfCrG8

#### 1. Your Information

First Name: ____________________
Family Name: ____________________
Initials: ____________________
E-mail address: ______________________________

Please state your institutional affiliation, academic position and postgraduate qualifications

______________________________________________________________

(As they would appear in a publication.)
(Refer to the publication policy of the study.)

Please indicate your role in this PEACH in Asia study project: (Check ALL that apply)
□ National Lead Investigator (National Coordinator)
□ Hospital Lead Investigator (Local Coordinator)

#### 2. Information of Your Institution

Country (or Region) name:

__________________________________________________

Full Name of Hospital:

__________________________________________________

Webpage (Homepage) Address of Hospital, if available

__________________________________________________

Level of Hospital Care (see definition below):
□ First level
□ Second level
□ Third level

**Table:** Definitions of Levels of Hospital Care

| Level of care | | Alternative terms commonly found in the literature |
|---|---|---|
| **First-Level** | Few specialties—mainly internal medicine, obstetrics and gynecology, pediatrics, and general surgery; often only one general practice physician or a nonphysician practitioner; limited laboratory services available for general but not specialized pathological analysis; from 50 to 250 beds. | Primary-level hospital<br>District hospital<br>Rural hospital<br>Community hospital<br>General hospital |
| **Second-Level** | More differentiated by function with as many as 5 to 10 clinical specialties; from 200 to 800 beds. | Regional hospital<br>Provincial hospital (or equivalent administrative area such as county)<br>General hospital |
| **Third-Level** | Highly specialized staff and technical equipment—for example, cardiology, intensive care unit, and specialized imaging units; clinical services highly differentiated by function; could have teaching activities; from 300 to 1,500 beds. | National hospital<br>Central hospital<br>Academic or teaching or university hospital |

Is this hospital:
□ Children's Hospital (a hospital that provides services exclusively to infants, children, adolescents, and young adults)

Is this hospital: (Check ALL that apply)
□ Government-Funded
□ Privately Funded
□ NGO/Mission/Charity Facility
□ University Hospital

Total Number of Hospital Beds: ________________________
Total Number of Operating Rooms: ________________________

Average Number of Surgical Cases per Month: ________________________
Average Number of Surgical Cases for Patients Under 16 Years per Month:
________________________

Number of Full-Time Specialist Anesthesiologists: ________________________

Do any of the specialist anesthesiologists handle pediatric cases (including neonatal cases) exclusively?

☐ Yes ☐ No

If yes, how many such pediatric specialist anesthesiologists are at your hospital?
_________________________________________

Is there a hospital protocol for pediatric pre-operative fasting?

☐ Yes ☐ No

If yes, is clear liquids allowed?

☐ More than 2 hours prior to anesthesia
☐ 2 hours prior to anesthesia
☐ 1 hour prior to anesthesia
☐ Less than 1 hour prior to anesthesia

Are blood transfusions performed at this hospital?

☐ Yes ☐ No

**Medication Availability**

Have you had **atropine** available every time you needed it?

☐ Always ☐ Sometimes ☐ Never

Have you had **epinephrine** (adrenaline) available every time you needed it?

☐ Always ☐ Sometimes ☐ Never

Have you had **fentanyl** available every time you needed it?

☐ Always ☐ Sometimes ☐ Never

Have you had **ketamine** available every time you needed it?

☐ Always ☐ Sometimes ☐ Never

**Equipment**

Do you have a reliable **electricity** supply?

☐ Always ☐ Sometimes ☐ Never

Do you have a reliable **oxygen** supply?
☐ Always ☐ Sometimes ☐ Never

Do you have functioning **incubators**?
☐ Yes ☐ No

Do you have functioning **electric patient warming devices**?
☐ Yes ☐ No

Is there a dedicated **Pediatric Emergency (Difficult) Airway Trolley** in the Operating Room?
☐ Yes ☐ No

Is there a hospital **protocol for pediatric emergency airway management**?
☐ Yes ☐ No

**Information on Local Ethical Approval**

Local Ethics Approval Confirmed
☐ Yes ☐ No
Name of Ethics Committee:

________________________________________

Other Local Regulatory Approval Confirmed
☐ Yes ☐ No
Name of Local Regulatory Committee:

________________________________________

I confirm that the information provided above is accurate.
☐ Yes

**Appendix: Publication Policy for the PEACH in Asia Study**

**Following the submission of the grant proposal, patient recruitment, data acquisition, cleaning, and analysis, authorship will be assigned based on contributions.**

1. Each participating institution with at least 5 patients can designate one collaborator to be mentioned in the publication.
2. For every additional 50 patients included, one additional collaborator can be designated.
3. These collaborators will be listed in the manuscript and traceable via PubMed.
4. Participating institutions may use their data upon request.
5. Proposals for secondary analyses can be submitted to the committee (the ASPA Executive Committee or the ASPA research special interest group) for approval. All papers resulting from the final analysis will be reviewed by the committee(s) before submission.
6. The sponsor of the study (ASPA research special interest group) may use anonymized data
7. for internal analyses and educational purposes.

**Abbreviations: ASPA = Asian Society of Pediatric Anesthesiologists**
