## Appendix 5 for "Protocol for the PEACH in Asia Study: A Prospective Multinational Multicenter Observational Study on the Epidemiology of Severe Critical Events in Pediatric Anesthesia in Asia"

**Appendix – Critical Events Definitions**

**(PEACH in Asia study)**

| | | |
|---|---|---|
| **1** | **Anaphylaxis** | The occurrence of any suspected IgE or non-IgE mediated severe allergic reaction leading to cardiovascular instability and/or severe bronchospasm and requiring immediate resuscitation (fluid resuscitation and epinephrine). |
| **2** | **Bronchospasm** | An increased respiratory effort, especially during expiration, and wheeze on auscultation.<br>If the patient is ventilated, bronchospasm may also be considered if a significant increase in peak inspiratory pressure (under volume controlled ventilation) or significant decrease in tidal volume (under pressure controlled ventilation) are observed. In all cases, any episode of airway constriction requiring the administration of a bronchodilator will be recorded. |
| **3** | **Cardiac Arrest (Peri-anesthetic)** | Cessation of circulation (e.g. pulseless electric activity, asystole, ventricular fibrillation/tachycardia) requiring open or closed chest compressions, or resulting in death, while the patient is in the care of the anesthetic team. |
| **4** | **Cardiovascular instability** | The occurrence of either one of the following:<br>1. **cardiac arrhythmia** defined as electrocardiogram(ECG) evidence of cardiac rhythm disturbance considered by clinical staff to be severe enough to require treatment (e.g. anti-arrhythmic agents, vasoactive agents, intravenous fluid, etc.). This includes arrhythmias occurring following regional analgesia and requiring intervention. For example: bradycardia requiring atropine, supraventricular tachycardia, atrial or ventricular tachyarrhythmia, torsade de pointe, etc.<br>2. **hypotension** defined as a drop in blood pressure requiring intervention by the anesthesiologist (fluid resuscitation and/or the administration of vasoactive drugs).<br>3. **bleeding** resulting in hypotension and necessitating unanticipated and unpredicted blood transfusion.<br>4. **cardiovascular instability** despite anticipated bleeding and transfusion (e.g.: liver transplant, scoliosis…). |

| | | |
|---|---|---|
| **5** | **Drug error** | The administration of a wrong drug, or a wrong dose given by any route, or a wrong site of administration, that has led to either respiratory/ cardiac/ neurological consequence or to an unplanned admission to the intensive care unit(ICU) or prolonged hospitalization. |
| **6** | **Laryngospasm** | Complete airway obstruction associated with rigidity of the abdominal and chest walls and leading to unsuccessful child's ventilation, or glottic closure associated with chest movement but silent unsuccessful child's respiratory efforts and assisted ventilation, unrelieved in both situations with simple jaw thrust and continuous positive airway pressure(CPAP) maneuvers and requiring the administration of medication (propofol, fentanyl, suxamethonium, rocuronium etc.) and/ or tracheal intubation. |
| **7** | **Neurological damage** | 1. In case of regional anaesthesia: the occurrence of nerve injury or spinal cord insult or seizure requiring resuscitation.<br>2. In case of general anaesthesia: any episode of seizure, pressures sore, episodes of loss of vision or new onset of central neurological impairment.<br>This includes peripheral nerve injury following positioning (ulnar nerve, external popliteal nerve) or puncture (median or ulnar nerve). |
| **8** | **Pulmonary Aspiration** | The presence of any non-respiratory secretions (bilious or particulate) in the airway as evidenced by laryngoscopy, suctioning, or bronchoscopy.<br>In a situation where there was suspicion of pulmonary aspiration but no positive aspiration of non-respiratory secretions, new clinical and/ or chest X-ray signs consistent with aspiration are accepted as evidence for it (e.g., new wheeze or crackles in the chest after regurgitation or vomiting incident). |
| **9** | **Stridor (Post-operative)** | A severe inspiratory flow limitation with sternal retraction, intrathoracic pressure swing, and potentially cyanosis occurring in the post-anesthesia care unit(PACU) and necessitating the administration of oxygen, intravenous steroids and/or epinephrine (nebulization) or tracheal intubation. |
