## Appendix 6 (REVISED in 2024) for "Protocol for the PEACH in Asia Study: A Prospective Multinational Multicenter Observational Study on the Epidemiology of Severe Critical Events in Pediatric Anesthesia in Asia"

**CRF 1: Preoperative Data (Before Anesthesia)**

| PATIENT INFORMATION & CONSENT | | |
|---|---|---|
| 1.1 | Country or Region Name | |
| | ***(choose single most appropriate)***<br>☐ 001: Afghanistan<br>☐ 002: Bahrain<br>☐ 003: Bangladesh<br>☐ 004: Bhutan<br>☐ 005: Brunei Darussalam<br>☐ 006: Cambodia<br>☐ 007: China<br>☐ 008: Democratic People's Republic of Korea<br>☐ 009: India<br>☐ 010: Indonesia<br>☐ 011: Iran (Islamic Republic of)<br>☐ 012: Iraq<br>☐ 013: Japan<br>☐ 014: Jordan<br>☐ 015: Kuwait<br>☐ 016: Lao People's Democratic Republic<br>☐ 017: Lebanon<br>☐ 018: Malaysia<br>☐ 019: Maldives | ☐ 020: Mongolia<br>☐ 021: Myanmar<br>☐ 022: Nepal<br>☐ 023: Oman<br>☐ 024: Pakistan<br>☐ 025: Philippines<br>☐ 026: Qatar<br>☐ 027: Republic of Korea<br>☐ 028: Saudi Arabia<br>☐ 029: Singapore<br>☐ 030: Sri Lanka<br>☐ 031: Syrian Arab Republic<br>☐ 032: Thailand<br>☐ 033: Timor-Leste<br>☐ 034: Turkey<br>☐ 035: United Arab Emirates<br>☐ 036: Viet Nam<br>☐ 037: Yemen<br>☐ 038: Hong Kong<br>☐ 039: Chinese Taipei |
| 1.2 | Hospital code | \|__\|__\|__\| (3 digit)<br>(Provided by the sponsor of this study) |
| 1.3 | Individual patient number at your hospital | \|__\|__\|__\| (3 digit)<br>(You can give a number in the order you collect the data in your hospital) |
| 2.1 | Was it necessary to obtain informed consent (IC) from guardians in order to include this patient in the study?<br>**(choose "No" if waived by local ethics committee or local research board)**<br>**(choose "No" if the IC was just necessary to provide clinical procedures such as surgery or anesthesia)** | ☐ Yes<br>☐ No |
| 2.2 | In case that IC was necessary for study enrollment, enter date when the IC was obtained | \|__\|__\|__\|__ \|-\|__\|__\|- \|__\|__\|<br>[ **YYYY – MM – DD** ]<br><br>☐ N/A |

| DEMOGRAPHICS | | |
|---|---|---|
| 3.1 | Date of Anesthesia / Sedation | \|__\|__\|__\|__ \|-\|__\|__\|- \|__\|__\|<br>[ **YYYY – MM – DD** ] |
| | 3.1.1 Anesthesia / Sedation time | ***(choose single most appropriate)***<br>☐ Opening hours of Operating room (i.e. Daytime of Weekdays)<br>☐ After hours/ Weekends/ Holidays |
| | 3.1.2 Duration of surgical/ non-surgical procedure | \|__\|__\|__\|__\| min [1-9999] |
| | 3.1.3 Duration of anesthesia/ sedation | \|__\|__\|__\|__\| min [1-9999] |
| 3.2 | Was the child premature?<br>(less than 37 weeks) | ☐ Yes<br>☐ No |
| | 3.2.1<br>If yes, gestational age at birth? | \|__\|__\| weeks [21-36]<br><br>☐ N/A |
| 3.3 | Age of the patient | ***(choose single most appropriate)***<br>☐ **Preterm neonatal:** The period at birth when a newborn is born before the full gestational period<br>☐ **from birth ≦27 days** (Term neonatal)<br>☐ **≧28 days, ≦12 months** (Infancy)<br>☐ **≧13 months, < 2 years** (Toddler)<br>☐ **≧2 years, ≦5 years** (Early childhood)<br>☐ **≧6 years, ≦11 years** (Middle childhood)<br>☐ **≧12 years, ≦15 years** (Early adolescence) |
| 3.4 | Sex of the patient | ☐ Male ☐ Female<br>☐ Unknown / Not yet determined after birth |
| 3.5 | Ethnicity of the patient | ***(choose single most appropriate)***<br>☐ Asian (e.g. Indian, Pakistani, Bangladeshi, Chinese, etc.)<br>☐ Arabic (North Africa, Middle East)<br>☐ Black (Caribbean, African)<br>☐ Spanish/Hispanic/Latino<br>☐ White<br>☐ Other |
| 3.6 | ASA Physical Status | ☐ 1 ☐ 2 ☐ 3 ☐ 4 ☐ 5<br>☐ 1E ☐ 2E ☐ 3E ☐ 4E ☐ 5E |
| 3.7 | Height of the patient | \|__\|__\|__\| cm [30-200]<br><br>☐ Not available |
| 3.8 | Weight of the patient | \|__\|__\|__\| . \|__\| kg [0.0-150.0]<br><br>☐ Not available<br><br>**(Please write numbers with 1 decimal places.)**<br>**(e.g. If the body weight of your patient is "15" kg, you need to write down "15.0" kg.)** |

| MEDICAL HISTORY | | |
|---|---|---|
| 4.1 | **Flu/cold:**<br>Does child currently (or in the 2 weeks preceding procedure) have flu or a cold? | ☐ Yes ☐ No ☐ Not available |
| 4.2 | **Wheezing/whistling:**<br>Has the child had wheezing or whistling spontaneously or after exercise in the last 12 months? | ☐ Yes ☐ No ☐ Not available |
| 4.3 | **Asthma:**<br>Has the child ever been diagnosed with asthma? | ☐ Yes ☐ No ☐ Not available |
| 4.4 | **Smoking:**<br>Does anyone in the family smoke? | ☐ Yes ☐ No ☐ Not available |
| 4.5 | **Allergy:**<br>Has the child ever had allergy? | ☐ Yes ☐ No ☐ Not available |
| | 4.5.1<br>**If yes, indicate all allergies that apply** | ☐ Not applicable<br><br>☐ Food ☐ Nut ☐ Latex<br>☐ Antibiotics ☐ Other |
| 4.6 | **Atopy:**<br>Is the child atopic? | ☐ Yes ☐ No ☐ Not available |
| 4.7 | **Fever** (exceeding 38.5℃ in the last 24 hours) | ☐ Yes ☐ No ☐ Not available |
| 4.8 | **Snoring:**<br>While sleeping, does the child snore? | ☐ Yes ☐ No ☐ Not available |
| 4.9 | **Medication:**<br>Does the child take any regular medication, natural products and/or homeopathic products? | ☐ Yes ☐ No ☐ Not available |
| 4.10 | **Handicap:**<br>Does the child have metabolic/ genetic disorder or neurological impairment? | ☐ Yes ☐ No ☐ Not available |
| 4.11 | **Anesthetic complication:**<br>Has the child had any previous documented anesthetic complication? | ☐ Yes ☐ No ☐ Not available |

| INDICATION | | |
|---|---|---|
| 5.1 | **Type of procedure** | ☐ **Surgical procedure**<br>☐ **Non- Surgical procedure** |
| | 5.1.1<br>**ONLY if "Surgical" in 5.1,**<br>indicate type of surgical procedure | ***(tick all that apply)***<br>☐ Cardiac surgery<br>☐ Cutaneous/Dermatology<br>☐ Ear-Nose-Throat<br>☐ Gastro/Abdominal/Hepato-biliary/Pancreas<br>☐ Head and Neck<br>☐ Neurosurgery<br>☐ Obstetric/ Gynecologic<br>☐ Ophthalmology<br>☐ Orthopedic<br>☐ Plastics (including cleft palate & lip)<br>☐ Thoracic<br>☐ Urological/Kidney<br><br>☐ Not applicable (i.e. Non-Surgical procedure) |
| | 5.1.2<br>**ONLY if "Non-Surgical procedure" in 5.1,**<br>indicate type of non-surgical procedure | ***(tick all that apply)***<br>☐ Biopsy<br>☐ Bone Marrow<br>☐ Bronchoscopy<br>☐ Burns dressing<br>☐ CT-Scan<br>☐ Dental<br>☐ Gastroenterology<br>☐ Lumbar puncture<br>☐ MRI (Magnetic Resonance Imaging)<br>☐ Ophthalmologic examination<br>☐ Venous access<br>☐ Other non-surgical<br><br>☐ Not applicable (i.e. Surgical procedure) |
| 5.3 | **Patient type** | ***(choose single most appropriate)***<br>☐ Outpatient ☐ Inpatient |

| ANAESTHESIA PLAN | | |
|---|---|---|
| 6.1 | **The senior anesthesiologist in charge**<br>What kind of Anesthesiologist? | ***(choose single most appropriate)***<br>☐ Specialist anesthesiologist with mainly pediatric practice (>50%)<br>☐ Specialist anesthesiologist with occasional pediatric anesthesia cases (<50%)<br>☐ Anesthesiologist in training<br>☐ Non-physician anesthesia provider (Nurse or Other Professionals) |
| | 6.1.1 **Experience**<br>For how many years the senior person in charge of the patient has been practicing? | \|__\|__\| yrs [0-60] |
| 6.2 | **Pre-Medications**<br>Was there any medication taken by the child just before the anesthesia? | ☐ Yes<br>☐ No |
| | 6.2.1<br>If yes, indicate medication(s): | ***(tick all that apply)***<br>☐ Not applicable (i.e. no pre-medications)<br><br>☐ Midazolam PO (oral)<br>☐ Midazolam IN (intra-nasal)<br>☐ Lorazepam IV (intra-venous)<br>☐ Temazepam PR (rectal)<br>☐ Temazepam PO (oral)<br>☐ Clonidine PO (oral)<br>☐ Clonidine IN (intra-nasal)<br>☐ Clonidine PR (rectal)<br>☐ Dexmedetomidine PO (oral)<br>☐ Dexmedetomidine IN (intra-nasal)<br>☐ Ketamine PO (oral)<br>☐ Ketamine IM (intra-muscular)<br>☐ Ketamine IV (intravenous)<br>☐ Chloral hydrate PO (oral)<br>☐ Chloral hydrate PR (rectal)<br>☐ Melatonin PO (oral)<br>☐ Acetaminophen (Paracetamol) PO (oral)<br>☐ Acetaminophen (Paracetamol) PR (rectal)<br>☐ NSAIDs (e.g. Ibuprofen) PO (oral)<br>☐ NSAIDs (e.g. Ibuprofen) PR (rectal)<br>☐ Local anesthetic cream/ tape (e.g. EMLA)<br>☐ Other |
| 6.3 | **Parental presence**<br>Was the child accompanied by<br>a parent or guardian during the induction? | ☐ Yes<br>☐ No |
| 6.4 | **Special monitoring**<br>Please specify | ***(tick all the apply)***<br>☐ Capnography<br>☐ Invasive arterial pressure monitoring<br>☐ Central venous pressure monitoring<br>☐ EEG derivated data (e.g. BIS)<br>☐ Near-infrared spectroscopy (NIRS)<br>☐ Neuromuscular monitoring (NMB monitoring) |

**CRF 2: Intraoperative Data (During anesthesia and 60 minutes afterwards)**

| INDUCTION | | |
|---|---|---|
| 7.1 | **Induction type at onset** | ***(Choose single most appropriate)***<br>☐ Inhalational<br>☐ Intravenous<br>☐ Intramuscular |
| 7.2 | **Rapid Sequence Induction** | ***(Choose single most appropriate)***<br>☐ Not applicable (i.e. Not rapid sequence induction)<br><br>☐ Yes: No mask ventilation<br>☐ Yes: Modified with mask ventilation |
| 7.3 | **Cricoid pressure** | ☐ Yes ☐ No |
| 7.4 | **Induction medication** | ***(tick all that apply)***<br>☐ Barbiturates (e.g. Thiopentone)<br>☐ Benzodiazepine (e.g. Midazolam, Remimazolam)<br>☐ Dexmedetomidine<br>☐ Etomidate<br>☐ Halothane<br>☐ Ketamine<br>☐ Nitrous Oxide<br>☐ Opioid(s)<br>☐ Propofol<br>☐ Sevoflurane<br>☐ Other |
| 7.5 | **Use of neuromuscular blocking agent(s) (NMBs) <u>at induction</u> and/ or <u>during maintenance</u>** | ***(tick all that apply)***<br>☐ No NMBs were used<br><br>☐ Atracurium<br>☐ Cis-atracurium<br>☐ Pancuronium<br>☐ Rocuronium<br>☐ Succinylcholine<br>☐ Vecuronium<br>☐ Other |
| | 7.5.2<br>**If NMBs used, reversal at the end?** | ***(choose single most appropriate)***<br>☐ Not applicable (i.e. No NMBs were used)<br><br>☐ Neostigmine<br>☐ Sugammadex<br>☐ The patient was extubated in the end in the OR, but NO reversals were used<br>☐ The patient was kept intubated after procedures |

| MAINTENANCE | | |
|---|---|---|
| 8.1 | **Anesthesia type during maintenance** | ***(choose single most appropriate)***<br>☐ Inhalational<br>☐ Total intravenous anesthesia (TIVA)<br>☐ Sedation (with NO AIRWAY DEVICES)<br>☐ Regional anesthesia alone |
| 8.2 | **Medication during maintenance** | ***(tick all that apply)***<br>☐ Barbiturates (e.g. Thiopentone)<br>☐ Benzodiazepines (e.g. Midazolam, Remimazolam)<br>☐ Desflurane<br>☐ Dexmedetomidine<br>☐ Halothane<br>☐ Isoflurane<br>☐ Ketamine<br>☐ Nitrous Oxide<br>☐ Opioid(s)<br>☐ Propofol<br>☐ Sevoflurane<br>☐ Other |
| 8.3 | **Regional anesthesia was provided by anesthesiologist** | ☐ Yes ☐ No |
| | 8.3.1<br>If “YES” in 8.3,<br>how was the regional anesthesia provided?<br>specify type | ***(Choose single most appropriate)***<br>☐ Not applicable (i.e. No regional anesthesia done)<br><br>☐ Landmarks<br>☐ Nerve stimulation (NS)<br>☐ Ultrasound (US) guided<br>☐ Combination of both NS and US |
| | 8.3.2<br>If “YES” in 8.3,<br>what type of regional anesthesia was provided?<br>specify type | ***(tick all that apply)***<br>☐ Not applicable (i.e. No regional anesthesia done)<br><br>☐ Caudal<br>☐ Craniofacial<br>☐ Epidural (except for Caudal)<br>☐ Ilioinguinal<br>☐ Intercostal<br>☐ Lower limb<br>☐ Paravertebral<br>☐ Penile<br>☐ Pudendal<br>☐ Rectus sheath (paraumblical)<br>☐ Spinal<br>☐ TAP (i.e. Transabdominal plane)<br>☐ Upper limb<br>☐ Other |

| AIRWAY MANAGEMENT | | |
|---|---|---|
| 9.1 | **Specify type of interface for airway management in the end**<br><br>e.g. In case that SGA was switched to ETT, please select ETT | ***(Choose single most appropriate)***<br>☐ Supraglottic airway (SGA) (e.g. LMA) → **If yes, please go to 9.2**<br>☐ Endotracheal tube (ETT) → **If yes, please go to 9.3**<br>☐ Anesthesia (Face) mask → **Skip 9.2 & 9.3**<br>☐ Non-invasive positive pressure ventilation (NPPV) → **Skip 9.2 & 9.3**<br>(e.g. nasal CPAP, high-flow nasal cannula)<br>☐ No airway devices (e.g. nasal cannula alone) → **Skip 9.2 & 9.3** |

| 9.2 | Only in case of SGA used | |
|---|---|---|
| | 9.2.1<br>**Insertion at** | ***(Choose single most appropriate)***<br>☐ 1st or 2nd attempt<br>☐ more than 3rd attempt |
| | 9.2.2<br>**Type of SGA** | ***(Choose single most appropriate)***<br>☐ Classic<br>☐ ProSeal<br>☐ Reinforced/ Flexible LMA<br>☐ Intubating LMA (ILMA)<br>☐ i-gel<br>☐ Other type |
| | 9.2.3<br>**Timing of removal of SGA** | ***(Choose single most appropriate)***<br>☐ Awake<br>☐ Semi-awake<br>☐ Deep anesthesia (Deep plane)<br>☐ SGA was not removed in the OR or in the PACU |

| 9.3 | Only in case of ETT used | |
|---|---|---|
| | 9.3.1<br>**Insertion at** | ***(Choose single most appropriate)***<br>☐ 1st or 2nd attempt<br>☐ more than 3rd attempt |
| | 9.3.2<br>**Type of ETT** | ***(Choose single most appropriate)***<br>☐ Micro-cuffed → Please go to 9.3.3<br>☐ Cuffed → Please go to 9.3.3<br>☐ Uncuffed → Skip the next question (9.3.3) then go to 9.3.4 |
| | 9.3.3<br>**If micro-cuffed or cuffed ETT was used, was cuff pressure monitored?** | ***(Choose single most appropriate)***<br>☐ Yes: cuff pressure was checked using a monometer<br>☐ No: cuff pressure was not checked using a manometer |
| | 9.3.4<br>**Intubation** | ***(Choose single most appropriate)***<br>☐ Direct laryngoscopy<br>☐ Video laryngoscopy<br>☐ Through Supraglottic Airway (SGA) Device<br>☐ Fiberoptic intubation<br>☐ Through tracheostomy<br>☐ Other |
| | 9.3.5<br>**Intubation way** | ***(Choose single most appropriate)***<br>☐ Oral<br>☐ Nasal<br>☐ Through tracheostomy |

| 9.3 | **Only in case of ETT used** | |
|---|---|---|
| | 9.3.6<br>**ETT type** | ***(Choose single most appropriate)***<br>☐ Classic (Normal)<br>☐ Micro-cuffed ETT<br>☐ Oral Ring–Adair–Elwyn (RAE) (“southpolar”)<br>☐ Nasal Ring–Adair–Elwyn (RAE) (“northpolar”)<br>☐ Reinforced (“Spiral”)<br>☐ Other |
| | 9.3.7<br>**Vocal Cords sprayed with lignocaine (lidocaine) prior to intubation?** | ☐ Yes<br>☐ No |
| | 9.3.8<br>**Cormack-Lehane score** | ***(Choose single most appropriate)***<br>☐ 1 ☐ 2 ☐ 3 ☐ 4 |
| | 9.3.9<br>**Timing of removal of ETT** | ***(Choose single most appropriate)***<br>☐ Awake<br>☐ Semi-awake<br>☐ Deep anesthesia (Deep plane)<br>☐ ETT was not removed in the OR or in the PACU |

| FLUIDS | | |
|---|---|---|
| 10 | **Was intravenous line secured for anesthesia / sedation?** | ☐ Yes<br>☐ No → **Please go to 11** |
| 10.1 | **Was glucose-containing fluid used?** | **☐ Yes → Please go to 10.1.2**<br>**☐ No → Please skip 10.1.2** |
| | 10.1.1<br>**If glucose-containing fluids was used, provide concentration** | ***(Choose single most appropriate)***<br>☐ 1%<br>☐ 2.5%<br>☐ 5%<br>☐ 10%<br>☐ Other % |
| 10.2 | **Was intravenous colloid used?** | ***(Choose single most appropriate)***<br>☐ No<br>☐ Albumin<br>☐ Synthetic colloids<br>☐ Other |
| 10.3 | **Was blood product used?** | ***(Choose single most appropriate)***<br>**☐ Yes → Please go to 10.3.1**<br>**☐ No → Please skip 10.3.1** |
| | 10.3.1<br>**If blood products were used, specify type of blood products** | ***(tick all that apply)***<br>☐ Packed red blood cells (pRBCs)<br>☐ Fresh frozen plasma (FFP)<br>☐ Platelets<br>☐ Fibrinogen<br>☐ Cryoprecipitate<br>☐ Other |

**CRF 3: Postoperative Data**
**Disposition after anesthesia/ sedation care**

| | | |
|---|---|---|
| 11.1 | Where was the patient transferred after the anesthesia/ sedation care? | ***(Choose single most appropriate)***<br>☐ Discharge from PACU (i.e. day-surgery)<br>☐ Ward<br>☐ Intensive care unit (ICU) |
| 11.2 | Oxygen delivery? | ***(Choose single most appropriate)***<br>☐ Yes, routinely<br>☐ Yes, if necessary (= as needed)<br>☐ No |

**11.1 Where was the patient transferred after the anesthesia /sedation care?**
In the choices, "Discharge from PACU (i.e. day-surgery)" defines "Discharge home from PACU." In the case who is admitted to ward from PACU, select the choice "Ward."
**11.2 Oxygen delivery?**
Some institutions or anesthesia practitioners give oxygen after general anesthesia "routinely" or "as a routine." In the case, kindly select the choice of "Yes, routinely", which is different from "Yes, if necessary" (= as needed).

**CRF 4: Perioperative Complications during anesthesia and up to 60 minutes afterwards**
**(As for the definitions of perioperative complications, please refer to the end of the case report form)**

**11.**
**Was any perioperative complications during anesthesia and up to 60 minutes afterwards?**

**☐ No Severe Critical Events**

1. Please skip the remaining questions. You have now completed all the questions!
2. Please enter the data directly into the electronic case report form (e-CRF) on the UMIN-INDICE cloud platform."

**☐ Yes → Please go to 12 for further details**

## 12.
### Please tick all the perioperative complications

- ☐ **1. Bronchospasm** → **section 13.1 for further details.**
- ☐ **2. Laryngospasm** → **section 13.2 for further details.**
- ☐ **3. Pulmonary aspiration** → **section 13.3 for further details.**
- ☐ **4. Drug error** → **section 13.4 for further details.**
- ☐ **5. Anaphylaxis** → **section 13.5 for further details.**
- ☐ **6. Cardiovascular instability** → **section 13.6 for further details**
- ☐ **7. Cardiac arrest** → **section 13.7 for further details**
- ☐ **8. Neurological damage** → **section 13.8 for further details**
- ☐ **9. Post-operative stridor** → **section 13.9 for further details**

| **13.1** | **Bronchospasm** | |
|---|---|---|
| 13.1.1 | **Bronchospasm, Time of occurrence?** ***(tick all that apply)*** | ☐ Induction<br>☐ Maintenance<br>☐ Awakening<br>☐ Post-anesthesia care unit (PACU) or Ward |
| 13.1.2 | **Bronchospasm, specify Treatment:** ***(tick all that apply)*** | ☐ Ventilation with intubation<br>☐ Bronchodilator<br>☐ Adrenaline (Epinephrine)<br>☐ Other |
| 13.1.3 | **Bronchospasm, Outcome of event?** ***(tick all that apply)*** | ☐ Uneventful<br>☐ Hypoxemia (SpO2 < 90% or 10% below the baseline)<br>☐ Prolonged intubation<br>☐ Cardiac arrest |

| **13.2** | **Laryngospasm** | |
|---|---|---|
| 13.2.1 | **Laryngospasm, Time of occurrence?** ***(tick all that apply)*** | ☐ Induction<br>☐ Maintenance<br>☐ Awakening<br>☐ Post-anesthesia care unit (PACU) or Ward |
| 13.2.2 | **Laryngospasm, specify Treatment:** ***(tick all that apply)*** | ☐ Ventilation with intubation<br>☐ CPAP with anesthesia (face) mask<br>☐ Propofol<br>☐ Opioid(s)<br>☐ Neuromuscular blockade(s)<br>☐ Other |
| 13.2.3 | **Laryngospasm, Outcome of event?** ***(tick all that apply)*** | ☐ Uneventful<br>☐ Hypoxemia (SpO2 < 90% or 10% below the baseline)<br>☐ Prolonged intubation<br>☐ Pulmonary edema<br>☐ Cardiac arrest |

| 13.3 | Pulmonary aspiration | |
|---|---|---|
| 13.3.1 | **Pulmonary aspiration, Time of occurrence?**<br>***(tick all that apply)*** | ☐ Induction<br>☐ Maintenance<br>☐ Awakening<br>☐ Post-anesthesia care unit (PACU) or Ward |
| 13.3.2 | **Pulmonary aspiration, specify Treatment:**<br>***(tick all that apply)*** | ☐ Broncho-tracheal suction<br>☐ Intubation<br>☐ CPAP<br>☐ Bronchodilator<br>☐ Other |
| 13.3.3 | **Pulmonary aspiration, Outcome of event?**<br>***(tick all that apply)*** | ☐ Uneventful<br>☐ Hypoxemia (SpO2 < 90% or 10% below the baseline)<br>☐ Prolonged intubation<br>☐ Cardiac arrest |

| 13.4 | Drug error | |
|---|---|---|
| 13.4.1 | **Drug error, Time of occurrence?**<br>***(tick all that apply)*** | ☐ Induction<br>☐ Maintenance<br>☐ Awakening<br>☐ Post-anesthesia care unit (PACU) or Ward |
| 13.4.2 | **Drug error, specify Type:**<br>***(tick all that apply)*** | ☐ Wrong dosage<br>☐ Wrong product<br>☐ Wrong site of administration<br>☐ Other |
| 13.4.3 | **Drug error, Treatment necessary?** | ☐ Yes ☐ No |
| 13.4.3 | **Drug error, Outcome of event?**<br>***(tick all that apply)*** | ☐ Uneventful<br>☐ Unplanned admission to ICU or Ward<br>☐ Cardiac arrest<br>☐ Other |

| 13.5 | Anaphylaxis | |
|---|---|---|
| 13.5.1 | **Anaphylaxis, Time of occurrence?**<br>***(tick all that apply)*** | ☐ Induction<br>☐ Maintenance<br>☐ Awakening<br>☐ Post-anesthesia care unit (PACU) or Ward |
| 13.5.2 | **Anaphylaxis, specify Treatment:**<br>***(tick all that apply)*** | ☐ Fluid resuscitation<br>☐ Intramuscular Adrenaline (Epinephrine)<br>☐ Intravenous Adrenaline (Epinephrine)<br>☐ Bronchodilator<br>☐ Intubation, followed by mechanical ventilation<br>☐ Cardiopulmonary resuscitation |
| 13.5.3 | **Anaphylaxis, Outcome of event?**<br>***(tick all that apply)*** | ☐ Uneventful<br>☐ Pulmonary edema<br>☐ Cardiac arrest<br>☐ Prolonged intubation |

| 13.6 | Cardiovascular instability | |
|---|---|---|
| 13.6.1 | **Cardiovascular instability, Time of occurrence?** ***(tick all that apply)*** | ☐ Induction<br>☐ Maintenance<br>☐ Awakening<br>☐ Post-anesthesia care unit (PACU) or Ward |
| 13.6.2 | **Cardiovascular instability, specify Type:** ***(tick all that apply)*** | ☐ Cardiac arrhythmia<br>☐ Hypotension<br>☐ Bleeding<br>☐ Other (e.g. Vasodilation) |
| 13.6.3 | **Cardiovascular instability, specify Treatment:** ***(tick all that apply)*** | ☐ Fluid resuscitation<br>☐ Blood product<br>☐ Vasopressor<br>☐ Atropine<br>☐ Antiarrhythmic drugs<br>☐ Defibrillation<br>☐ Electrical cardioversion<br>☐ Other |
| 13.6.4 | **Cardiovascular instability, Outcome of event?** ***(tick all that apply)*** | ☐ Uneventful<br>☐ Coagulopathy<br>☐ Cardiac arrest<br>☐ Unplanned admission to ICU or Ward |

| 13.7 | Cardiac arrest | |
|---|---|---|
| 13.8.1 | **Cardiac arrest, Time of occurrence?** ***(tick all that apply)*** | ☐ Induction<br>☐ Maintenance<br>☐ Awakening<br>☐ Post-anesthesia care unit (PACU) or Ward |
| 13.8.2 | **Cardiac arrest, specify Treatment:** ***(tick all that apply)*** | ☐ Closed chest compression<br>☐ Open (Direct) cardiac massage<br>☐ Defibrillation<br>☐ Adrenaline (Epinephrine)<br>☐ Extracorporeal membrane oxygenation (i.e. E-CPR) |
| 13.8.3 | **Cardiac arrest, Outcome of event?** ***(tick all that apply)*** | ☐ Uneventful<br>☐ Unplanned admission to ICU or Ward<br>☐ Poor neurological outcome<br>☐ Death |

| 13.8 | Neurological damage(s) | |
|---|---|---|
| 13.8.1 | **Neurological damage, Time of occurrence?** ***(tick all that apply)*** | ☐ Induction<br>☐ Maintenance<br>☐ Awakening<br>☐ Post-anesthesia care unit (PACU) or Ward |
| 13.8.2 | **Neurological damage, Treatment necessary?** | ☐ Yes ☐ No |
| 13.8.3 | **Anaphylaxis, Outcome of event?** ***(tick all that apply)*** | ☐ Uneventful<br>☐ Unplanned admission to ICU or Ward<br>☐ Poor neurological outcome<br>☐ Death |

| **13.9** | **Postoperative stridor** | |
|---|---|---|
| 13.9.1 | **Stridor, Time of occurrence?** ***(tick all that apply)*** | ☐ Awakening<br>☐ Post-anesthesia care unit (PACU) or Ward |
| 13.9.2 | **Stridor, specify Treatment:** ***(tick all that apply)*** | ☐ CPAP<br>☐ Adrenaline (Epinephrine)<br>☐ Other |
| 13.9.3 | **Stridor, Outcome of event?** ***(tick all that apply)*** | ☐ Uneventful<br>☐ Unplanned admission to ICU or Ward<br>☐ Intubation |

**Thank you for completing the data collection process!**

1. Please enter all data directly into the electronic case report form (e-CRF) on the cloud platform (UMIN-INDICE).
2. If you encounter any difficulties, please contact your national coordinator or the principal investigator for assistance.

**Severe Critical Events Definitions (PEACH in Asia study)**

| | | |
|---|---|---|
| **1** | **Bronchospasm** | An increased respiratory effort, especially during expiration, and wheeze on auscultation.<br>If the patient is ventilated, bronchospasm may also be considered if a significant increase in peak inspiratory pressure (under volume controlled ventilation) or significant decrease in tidal volume (under pressure controlled ventilation) are observed.<br>In all cases, any episode of airway constriction requiring the administration of a bronchodilator will be recorded. |
| **2** | **Laryngospasm** | Complete airway obstruction associated with rigidity of the abdominal and chest walls and leading to unsuccessful child's ventilation, or glottic closure associated with chest movement but silent unsuccessful child's respiratory efforts and assisted ventilation, unrelieved in both situations with simple jaw thrust and continuous positive airway pressure(CPAP) maneuvers and requiring the administration of medication (propofol, fentanyl, suxamethonium, rocuronium etc.) and/ or tracheal intubation. |
| **3** | **Pulmonary aspiration** | The presence of any non-respiratory secretions (bilious or particulate) in the airway as evidenced by laryngoscopy, suctioning, or bronchoscopy.<br>In a situation where there was suspicion of pulmonary aspiration but no positive aspiration of non-respiratory secretions, new clinical and/ or chest X-ray signs consistent with aspiration are accepted as evidence for it (e.g., new wheeze or crackles in the chest after regurgitation or vomiting incident). |
| **4** | **Drug error** | The administration of a wrong drug, or a wrong dose given by any route, or a wrong site of administration, that has led to either respiratory/ cardiac/ neurological consequence or to an unplanned admission to the intensive care unit(ICU) or prolonged hospitalization. |
| **5** | **Anaphylaxis** | The occurrence of any suspected IgE or non-IgE mediated severe allergic reaction leading to cardiovascular instability and/or severe bronchospasm and requiring immediate resuscitation (fluid resuscitation and epinephrine). |
| **6** | **Cardiovascular instability** | The occurrence of either one of the following:<br>1. **cardiac arrhythmia** defined as electrocardiogram(ECG) evidence of cardiac rhythm disturbance considered by clinical staff to be severe enough to require treatment (e.g. anti-arrhythmic agents, vasoactive agents, intravenous fluid, etc.). This includes arrhythmias occurring following regional analgesia and requiring intervention. For example: bradycardia requiring atropine, supraventricular tachycardia, atrial or ventricular tachyarrhythmia, torsade de pointe, etc.<br>2. **hypotension** defined as a drop in blood pressure requiring intervention by the anesthesiologist (fluid resuscitation and/or the administration of vasoactive drugs).<br>3. **bleeding** resulting in hypotension and necessitating unanticipated and unpredicted blood transfusion.<br>4. **cardiovascular instability** despite anticipated bleeding and transfusion (e.g.: liver transplant, scoliosis…). |
| **7** | **Cardiac Arrest** | Cessation of circulation (e.g. pulseless electric activity, asystole, ventricular fibrillation/tachycardia) requiring open or closed chest compressions, or resulting in death, while the patient is in the care of the anesthetic team. |
| **8** | **Neurological damage** | 1. In case of regional anaesthesia: the occurrence of nerve injury or spinal cord insult or seizure requiring resuscitation.<br>2. In case of general anaesthesia: any episode of seizure, pressures sore, episodes of loss of vision or new onset of central neurological impairment.<br>This includes peripheral nerve injury following positioning (ulnar nerve, external popliteal nerve) or puncture (median or ulnar nerve). |
| **9** | **Stridor (Post-operative)** | A severe inspiratory flow limitation with sternal retraction, intrathoracic pressure swing, and potentially cyanosis occurring in the post-anesthesia care unit(PACU) and necessitating the administration of oxygen, intravenous steroids and/or epinephrine (nebulization) or tracheal intubation. |
