## Appendix 7 for "Protocol for the PEACH in Asia Study: A Prospective Multinational Multicenter Observational Study on the Epidemiology of Severe Critical Events in Pediatric Anesthesia in Asia"

### Correspondence Table form for PEACH in Asia study

| **BASIC INFORMATION** | |
|---|---|
| **COUNTRY OR REGION** | Code |
| | Name |
| **NATIONAL CO-ORDINATOR** | Name |
| | Contact e-mail address |
| **INSTITUTION** | Code |
| | Name |
| | Address |
| **LOCAL CO-ORDINATOR** | Name |
| | Affiliation |
| | Contact e-mail address |

Information on anesthesia, sedation, and procedures provided at the participating institution and patient information (age, height, weight, etc.) necessary to investigate their effects and risks are registered.

Study identification number (i.e. study ID) not related to patients' personal information are used in registering those information

***This correspondence table linking study ID and patients' institutional ID is strictly controlled in the participating institution and should not be provided to the sponsor of the study, the ASPA research committee.***

**Correspondence Table linking Study ID and Patients' ID at the participating institution**
**(October 31st, 2022: created by Soichiro Obara)**

**Country or Region Code:** ___________________

**Institution Code:** __________________________

| Study ID at your institution | Patient ID at your institution |
|---|---|
| 001 | |
| 002 | |
| 003 | |
| 004 | |
| 005 | |
| 006 | |
| 007 | |
| 008 | |
| 009 | |
| 010 | |
| 011 | |
| 012 | |
| 013 | |
| 014 | |
| 015 | |
| 016 | |
| 017 | |
| 018 | |
| 019 | |
| 020 | |
| 021 | |
| 022 | |
| 023 | |
| 024 | |
| 025 | |
| 026 | |
| 027 | |
| 028 | |
| 029 | |
| 030 | |
| 031 | |
| 032 | |
| 033 | |
| 034 | |
| 035 | |
| 036 | |
| 037 | |
| 038 | |
| 039 | |
| 040 | |
| 041 | |
| 042 | |
| 043 | |
| 044 | |
| 045 | |
| 046 | |
| 047 | |
| 048 | |
| 049 | |
| 050 | |
| 051 | |
| 052 | |
| 053 | |
| 054 | |
| 055 | |
| 056 | |
| 057 | |
| 058 | |
| 059 | |
| 060 | |

**Correspondence Table linking Study ID and Patients' ID at the participating institution**
**(October 31st, 2022: created by Soichiro Obara)**

**Country or Region Code:** ___________________

**Institution Code:** __________________________

| Study ID at your institution | Patient ID at your institution |
|---|---|
| 061 | |
| 062 | |
| 063 | |
| 064 | |
| 065 | |
| 066 | |
| 067 | |
| 068 | |
| 069 | |
| 070 | |
| 071 | |
| 072 | |
| 073 | |
| 074 | |
| 075 | |
| 076 | |
| 077 | |
| 078 | |
| 079 | |
| 080 | |
| 081 | |
| 082 | |
| 083 | |
| 084 | |
| 085 | |
| 086 | |
| 087 | |
| 088 | |
| 089 | |
| 090 | |
| 091 | |
| 092 | |
| 093 | |
| 094 | |
| 095 | |
| 096 | |
| 097 | |
| 098 | |
| 099 | |
| 100 | |
| 101 | |
| 102 | |
| 103 | |
| 104 | |
| 105 | |
| 106 | |
| 107 | |
| 108 | |
| 109 | |
| 110 | |
| 111 | |
| 112 | |
| 113 | |
| 114 | |
| 115 | |
| 116 | |
| 117 | |
| 118 | |
| 119 | |
| 120 | |

**Correspondence Table linking Study ID and Patients' ID at the participating institution (October 31st, 2022: created by Soichiro Obara)**

Country or Region Code: ___________________

Institution Code: __________________________

| Study ID at your institution | Patient ID at your institution |
|---|---|
| 121 | |
| 122 | |
| 123 | |
| 124 | |
| 125 | |
| 126 | |
| 127 | |
| 128 | |
| 129 | |
| 130 | |
| 131 | |
| 132 | |
| 133 | |
| 134 | |
| 135 | |
| 136 | |
| 137 | |
| 138 | |
| 139 | |
| 140 | |
| 141 | |
| 142 | |
| 143 | |
| 144 | |
| 145 | |
| 146 | |
| 147 | |
| 148 | |
| 149 | |
| 150 | |
| 151 | |
| 152 | |
| 153 | |
| 154 | |
| 155 | |
| 156 | |
| 157 | |
| 158 | |
| 159 | |
| 160 | |
| 161 | |
| 162 | |
| 163 | |
| 164 | |
| 165 | |
| 166 | |
| 167 | |
| 168 | |
| 169 | |
| 170 | |
| 171 | |
| 172 | |
| 173 | |
| 174 | |
| 175 | |
| 176 | |
| 177 | |
| 178 | |
| 179 | |
| 180 | |

**Correspondence Table linking Study ID and Patients' ID at the participating institution**
**(October 31st, 2022: created by Soichiro Obara)**

**Country or Region Code:** ___________________

**Institution Code:** __________________________

| Study ID at your institution | Patient ID at your institution |
|---|---|
| 181 | |
| 182 | |
| 183 | |
| 184 | |
| 185 | |
| 186 | |
| 187 | |
| 188 | |
| 189 | |
| 190 | |
| 191 | |
| 192 | |
| 193 | |
| 194 | |
| 195 | |
| 196 | |
| 197 | |
| 198 | |
| 199 | |
| 200 | |
| 201 | |
| 202 | |
| 203 | |
| 204 | |
| 205 | |
| 206 | |
| 207 | |
| 208 | |
| 209 | |
| 210 | |
| 211 | |
| 212 | |
| 213 | |
| 214 | |
| 215 | |
| 216 | |
| 217 | |
| 218 | |
| 219 | |
| 220 | |
| 221 | |
| 222 | |
| 223 | |
| 224 | |
| 225 | |
| 226 | |
| 227 | |
| 228 | |
| 229 | |
| 230 | |
| 231 | |
| 232 | |
| 233 | |
| 234 | |
| 235 | |
| 236 | |
| 237 | |
| 238 | |
| 239 | |
| 240 | |

**Correspondence Table linking Study ID and Patients' ID at the participating institution**
**(October 31st, 2022: created by Soichiro Obara)**

**Country or Region Code:** ___________________

**Institution Code:** ___________________________

| Study ID at your institution | Patient ID at your institution |
|---|---|

**Correspondence Table linking Study ID and Patients' ID at the participating institution**
**(October 31st, 2022: created by Soichiro Obara)**

**Country or Region Code:** ____________________

**Institution Code:** ____________________________

| Study ID at your institution | Patient ID at your institution |
|---|---|

**Correspondence Table linking Study ID and Patients' ID at the participating institution**
**(October 31st, 2022: created by Soichiro Obara)**

**Country or Region Code:** ___________________

**Institution Code:** ___________________________

| Study ID at your institution | Patient ID at your institution |
|---|---|

**Correspondence Table linking Study ID and Patients' ID at the participating institution**
**(October 31st, 2022: created by Soichiro Obara)**

**Country or Region Code:** ___________________

**Institution Code:** __________________________

| Study ID at your institution | Patient ID at your institution |
|---|---|

**Correspondence Table linking Study ID and Patients' ID at the participating institution (October 31st, 2022: created by Soichiro Obara)**

**Country or Region Code:** ___________________

**Institution Code:** __________________________

| Study ID at your institution | Patient ID at your institution |
|---|---|
