## Appendix 8 for "Protocol for the PEACH in Asia Study: A Prospective Multinational Multicenter Observational Study on the Epidemiology of Severe Critical Events in Pediatric Anesthesia in Asia"

### Notification of IRB Review

#### Protocol #: 2021-52

Date: November 30, 2021

To: Soichiro Obara, MD, DrPH
Department of Anesthesia, Tokyo Metropolitan Ohtsuka Hospital
Graduate School of Public Health, Teikyo University

From: Institutional Review Board at Tokyo Metropolitan Ohtsuka Hospital
2-8-1, Minamiohtsuka, Toshima-ku, Tokyo, 170-8476, Japan

**Title of Clinical Research:**
Peri-anesthetic morbidity in children in Asia: a prospective multinational multicenter observational study to investigate epidemiology of severe critical events in pediatric anesthesia in Asia
(PEACH in Asia: PEri-Anesthetic morbidity in CHildren in Asia)

| | |
|---|---|
| **IRB Review Type:** | Expedited |
| **IRB Approval Date:** | November 30th, 2021 |
| **Approval/Activation Date:** | November 30th, 2021 |
| **Clinical Trial Registration:** | UMIN Clinical Trials Registry (UMIN-CTR) # UMIN000046328 |

The IRB has determined as it follows:

The proposed study is an observational study with no interference with the child's routine care, which is in accordance to the good clinical practice and no research-related interventions, will be introduced. Therefore, no ethical concerns are expected and ethical approval may not be required in some centers. However, where ethical approval is required, this approval must be obtained before the start of inclusion.
In all cases, all participating centers must submit the study to the local Institutional Review Board for ethical judgment and obtain document of proof that the trial has been subject to IRB/IEC review and given approval/favorable opinion and/or waive the consent. If informed consent is not required by the local IRB, a written waiver must be obtained from the Institutional Review Board. This process should take place prior to initiation of the trial and in compliance with the applicable national regulatory requirement(s).

As Principal Investigator, you are responsible for the following:
1. Ensuring that this project is conducted in compliance with this determination.
2. Submission of significant proposed changes to this project to ensure that the project continues to meet the criteria for exemption.
