## Appendix 9 for "Protocol for the PEACH in Asia Study: A Prospective Multinational Multicenter Observational Study on the Epidemiology of Severe Critical Events in Pediatric Anesthesia in Asia"

Regarding PEACH in Asia study data capturing system
By Soichiro Obara on November 16th, 2022

**November 16th, 2022**

### RE: the PEACH in Asia study data capturing system, UMIN-INDICE system

To Whom It May Concern:

I am writing to give further information about the secure online UMIN-INDICE (Internet Data and Information Center for Medical Research) database which will be used to collect data for the PEACH in Asia (Peri-anesthetic morbidity in children in Asia: a prospective multinational multicenter observational study to investigate epidemiology of severe critical events in pediatric anesthesia in Asia) study.

UMIN (University hospital Medicine Information Network) was established in 1989 as a cooperative organization for national medical schools in Japan, sponsored by the Ministry of Education, Culture, Science, Sports and Technology (MEXT), Japan. Its most services are now made available to other health care researchers via the Internet. UMIN Center is an organization which provides a network composed of the 42 national University Hospital of Japan. The Center is located in the University of Tokyo Hospital and provides services to the whole Japan.

**UMIN website (in English): https://www.umin.ac.jp/english/whatisumin.htm**

UMIN is now the largest and most versatile academic network information center for biomedical sciences in the world, and it is now considered as indispensable information infrastructure for the Japanese medical community.

The PEACH in Asia study has been registered with the ICMJE (International Committee of Medical Journal Editors)-approved registry, the UMIN-Clinical Trial Registry (UMIN-CTRCTR) as it follows:
**The clinical registry ID: UMIN000046328**
The webpage:
**https://center6.umin.ac.jp/cgi-open-bin/ctr_e/ctr_view.cgi?recptno=R000052803**

UMIN-INDICE is the service to register and collect randomized cases of the clinical epidemiologic studies which researchers lead as scientific studies. UMIN-INDICE databases in the UMIN center have been successfully used for a number of international studies. The UMIN-INDICE database used for the PEACH in Asia study is run by the UMIN center which is physically secured.

Regarding PEACH in Asia study data capturing system
By Soichiro Obara on November 16th, 2022

The security of the study UMIN-INDICE database system is governed by the policies of the UMIN center in Japan, in accordance with the requirements of the General Data Protection Regulations (GDPR).

The study will be conducted at collaborating sites in accordance with the country-specific data protection requirements. Once data collection is complete, the electronic research files containing anonymized data will be stored on secure non-networked desktop computers for up to 5 years, in line with current regulations. Access will be restricted to the research sponsors, the Asian Society of Paediatric Anaesthesiologists (ASPA) research committee. Personal data will remain securely at each local participating hospital.

No sensitive or identifiable data will be collected on the database; the patient's clinical team will only upload non-identifiable data. Access to data will be restricted, each individual local collaborator entering data for the PEACH in Asia study will have their own username and password. Each patient will be allocated a unique study number at entry. The central research team, the ASPA research committee members including the principal investigator (Soichiro Obara) will not have any access to patient identifiable data. All communication will use this study number. All data will be analyzed and reported in summary format. No individual patient data will be identifiable in public reports.

Yours faithfully,

**Soichiro Obara, MD DrPH**

Soichiro Obara

**On behalf of the PEACH in Asia study steering committee and the Asian Society of Paediatric Anaesthesiologists (ASPA) research committee**

Department of Anesthesia, Tokyo Metropolitan Ohtsuka Hospital, Tokyo, Japan
Address: 2-8-1, Minamiotsuka, Toshima-ku, Tokyo 170-8476
